# Kidney Shape Statistical Analysis: Associations with Disease and Anthropometric Factors

**DOI:** 10.1101/2023.06.26.23291901

**Authors:** Marjola Thanaj, Nicolas Basty, Madeleine Cule, Elena P Sorokin, Brandon Whitcher, Ramprakash Srinivasan, Rachel Lennon, Jimmy D Bell, E Louise Thomas

## Abstract

**Background:** Organ measurements derived from magnetic resonance imaging (MRI) have the potential to enhance our understanding of the precise phenotypic variations underlying many clinical conditions.

**Methods:** We applied morphometric methods to study the kidneys by constructing surface meshes from kidney segmentations from abdominal MRI data in 38,868 participants in the UK Biobank. Using mesh-based analysis techniques based on statistical parametric maps (SPMs), we were able to detect variations in specific regions of the kidney and associate those with anthropometric traits as well as disease states including chronic kidney disease (CKD), type-2 diabetes (T2D), and hypertension. Statistical shape analysis (SSA) based on principal component analysis was also used within the disease population and the principal component scores were used to assess the risk of disease events.

**Results:** We show that CKD, T2D and hypertension were associated with kidney shape. Age was associated with kidney shape consistently across disease groups. Body mass index (BMI) and waist-to-hip ratio (WHR) were also associated with kidney shape for the participants with T2D. Using SSA, we were able to capture kidney shape variations, relative to size, angle, straightness, width, length, and thickness of the kidneys, within disease populations. We identified significant associations between both left and right kidney length and width and incidence of CKD (hazard ratio (HR): 0.74, 95% CI: 0.61-0.90, p<0.05, in the left kidney; HR: 0.76, 95% CI: 0.63-0.92, p<0.05, in the right kidney) and hypertension (HR: 1.16, 95% CI: 1.03-1.29, p<0.05, in the left kidney; HR: 0.87, 95% CI: 0.79-0.96, p<0.05, in the right kidney).

**Conclusions:** The results suggest that shape-based analysis of the kidneys can augment studies aiming at the better categorisation of pathologies associated with acute and chronic kidney conditions.

## Introduction

The incidence of conditions such as chronic kidney disease (CKD), type-2 diabetes (T2D), and hypertension are rising and are amongst the leading causes of death globally [1]. The prevalence of CKD, which increases with age, is significantly common among older people, with an adverse effect on longevity [2]. Progression of CKD is generally tracked as a gradual decline in glomerular filtration rate (GFR), however there are suggestions that changes in kidney volume occur much earlier in the disease process and can accurately predict disease [3, 4]. Changes in kidney volume, as well as kidney length and structure, have additionally been reported in metabolic diseases including obesity, T2D, and hypertension, these act as drivers of CKD progression reducing kidney function [5–7].

Recently the automated segmentation and measurement of kidney volume from magnetic resonance imaging (MRI) has become more commonplace, enabling rapid measurement and the ability to obtain detailed anatomical information [8]. While this advance has enhanced our understanding of the kidney at a population level, additional knowledge regarding morphological changes and regional variation in response to particular conditions are still lacking.

Three-dimensional (3D) mesh-derived phenotypes capture additional information related to morphological and regional organ variation using statistical parametric maps (SPMs) and may be used to map more subtle differences between a healthy and diseased state. A similar approach is statistical shape analysis (SSA), which can be used to transform the spatially correlated data into a smaller number of principal components and characterise variations in organ shape across a population. These morphometric analyses offer a way to model the human body non-invasively and have been widely used to model bones [9, 10], abdominal organs [11–13], the brain [14, 15], the heart [16, 17] and the aorta [18, 19]. However, they have been less frequently applied to abdominal organs, despite known morphological changes occurring in clinical conditions [3].

In the current study, we have applied computational image analysis to identify factors associated with variation in kidney shape in a region-specific manner and assessed whether this can be used to identify morphological variation associated with CKD, T2D, and hypertension. We further investigated whether the emerging 3D kidney mesh-derived phenotypes can add to the prediction of disease outcomes.

## Methods

### Data

Full details regarding the UK Biobank abdominal MRI acquisition protocol have previously been reported [20]. The data included here focused on the neck-to-knee Dixon MRI acquisitions. All data were processed and segmented using automated methods [8].

Participant data from the UK Biobank cohort was obtained through UK Biobank Access Application number 44584. The UK Biobank has approval from the North West Multi-Centre Research Ethics Committee (REC reference: 11/NW/0382). All methods were performed in accordance with the relevant guidelines and regulations, and informed consent was obtained from all participants. Researchers may apply to use the UKBB data resource by submitting a health-related research proposal that is in the public interest. Additional information may be found on the UK Biobank researchers and resource catalogue pages (www.ukbiobank.ac.uk).

### Quality Control

Participants with missing clinical, anthropometric, or biochemical data, as well as those with Dixon MRI datasets that did not have full anatomical coverage, were excluded from the study (i.e organs with zero volumes). We further performed quality control by visually inspecting potential outliers in the 3D kidney mesh-derived phenotype (i.e. extremely high values). To ensure comprehensive anatomical coverage, we also discarded kidney segmentations with volumes less than 30 ml from our analysis. For consistency in the sample size, participants with missing data or segmentations below the lower limit threshold for one kidney were excluded from the study even if the other kidney had full coverage. Overall, from the initial 44,455 participants, data from of 5,587 participants did not pass quality control and were excluded from the final analysis (12.6% of the dataset excluded, of which 47% were male, 64.7 ± 7.8 years old, with a BMI 26.3 ± 4.5 kg/m^2^ [mean ± standard deviation (SD)]), leaving a final dataset of 38,868 participants.

### Study Design

#### Phenotype Definitions

Anthropometric measurements including age, body mass index (BMI), waist and hip circumferences, and systolic and diastolic blood pressure were taken at the UK Biobank imaging visit, and ethnicity was defined based on the self-reported ethnic background at the initial assessment visit (field: 21000). For the purpose of our analysis we categorised ethnic background as follows: 0 for “White” and 1 for any other non-white ethnic background (due to small numbers of non-white participants in this dataset (3.1%)). Sex was self-reported and included those recorded by the NHS and those obtained at the initial assessment visit (field: 31).

Biological samples for serum creatinine (field: 30700) were measured in *millimole*/*L*, sodium in urine (field: 30530) in *millimole*/*L* and urea (field: 30670) in *millimole*/*L* units were taken on the initial assessment visit. Estimated glomerular filtration rate (eGFR) was calculated based on the CKD-EPI creatinine equation (2009) [21] as follows:

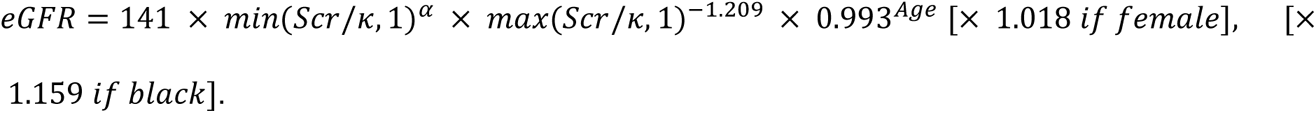

where *Scr* is serum creatinine in converted into *mg*/*dL* units, κ is 0.7 for females and 0.9 for males, *α* is −0.329 for females and −0.411 for males, *min* is the minimum of Scr/κ or 1, and *max* is the maximum of Scr/κ or 1. In the aforementioned equation, ethnicity for “Black” was defined based on the continental genetic ancestry for “African” ancestry (https://pan.ukbb.broadinstitute.org) and if missing from the self-reported ethnic background (field: 21000) for “Black or Black British”.

Questionnaire information from the UK Biobank imaging visit was used to determine alcohol intake frequency (field: 1558), smoking status (field: 20116), and usage of ibuprofen medication (field: 6154, field: 20003). For our analysis, we categorised alcohol intake frequency as 1 for “Daily or almost daily” and 0 otherwise and smoking status as 1 for “Current” and 0 for “Previous” and “Never”. It should be noted that the UK Biobank initial assessment visit preceded the imaging visit by 9 ± 1.7 years.

#### Association between mesh-derived phenotypes and disease

To assess the associations between the 3D mesh-derived phenotype, and the anthropometric covariates, we analysed the data of 38,868 UK Biobank participants.

We selected diseases known to be associated with kidney health, and those previously associated with changes in kidney volume [8]. In part, this was also guided by the number of patients available in the UK Biobank with diseases of interest. We included participants with chronic kidney disease (CKD), and T2D as well as participants with hypertension (see Disease Definitions in supporting information and supplementary Table S1).

### Image Registration and Mesh Construction

The process for organ template construction has been previously detailed in [22, 23]. Here, we constructed a template using the kidney segmentations from a sex-balanced European ancestry cohort of 200 participants. The characteristics of the template population are provided in Supplementary Table S2. We then constructed 3D surface meshes from the template image and all participants’ segmentations using the marching cubes algorithm and smoothed using a Laplacian filter [24]. Supplementary Fig. S1 illustrates a brief diagram for the construction of average kidney template meshes.

The registration process has been previously detailed in [23]. In brief, we used rigid registration to remove the position and orientation difference between all participant-specific surfaces and template surfaces. Then we registered the template to the participant’s segmentations via affine and non-rigid registration. The template mesh was then propagated to each participant mesh using the deformation fields obtained from the non-rigid registration. Hence, all surface meshes are parameterised with the same number of vertices (approximately 4,000) ensuring each vertex was anatomically accurate and consistent across all participants while preserving the size and shape information for subsequent analysis. All the steps for the template-to-subject registration were performed using the Image Registration Toolkit (IRTK) (https://biomedia.doc.ic.ac.uk/software/irtk).

To determine the regional outward or inward adaptations in kidney surface in comparison to an average kidney shape for each participant, the surface-to-surface (S2S) distance was measured by computing the signed distance between the template surface and the participants’ surface mesh at each vertex [23].

### Mass Univariate Regression Analysis

Associations between S2S distances and anthropometric variables were estimated using a linear regression framework. The linear regression model is expressed as follows:

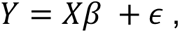

where, Y is a *n*_*s*_ × *n*_*v*_ matrix containing *n*_*s*_ subjects from a sample of the population under study and *n*_*v*_ is the number of voxels in the mesh, X is the *n*_*s*_ × p design matrix of p known covariates (including the intercept) and the relevant variables for each subject. X is related to Y by the vector of the estimated regression coefficients 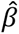 . Finally ϵ is a *n*_*s*_ × *n*_*v*_ matrix which is independent and identically distributed across the subjects and is assumed to be a zero-mean Gaussian process [25]. We applied threshold-free cluster enhancement (TFCE) [26] and permutation testing to assess the associations between S2S distances and anthropometric covariates, adjusted for relevant covariates with the correction to control the false discovery rate (FDR), as previously described [22].

Specifically, we performed an SPM framework, mass univariate regression (MUR) analysis using a refined version of the R package *mutools3D* [27] adjusted for multiple comparisons by applying the FDR procedure using the Benjamini-Hochberg method [28] to all the TFCE-derived p-values for each vertex and each model using 1,000 permutations. The estimated regression coefficients 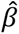 for each of the relevant covariates and their related TFCE-derived p-values after correction for multiple testing, were then displayed at each vertex in the mesh on the whole 3D kidney anatomy, providing the spatially-distributed associations. The MUR model for deriving associations between clinical parameters and a 3D phenotype is outlined in supplementary Fig. S2.

To determine which factors were associated with kidney shape and size, we fitted a linear regression model for each vertex with age, sex, ethnicity, BMI, waist-to-hip ratio (WHR), alcohol drinker status, smoking status, ibuprofen medication, sodium in urine, urea, CKD, T2D, and hypertension as predictors and applied a correction to control the FDR. It is well known that the number of nephrons decreases with age which can contribute to the decline in kidney function [4]. Hence, to investigate whether there was a stronger relationship in terms of accelerating change between age and CKD, T2D, and hypertension we included interaction terms between age and all clinical conditions. We then included interaction terms between BMI and T2D, and between WHR and T2D, as markers of obesity, to investigate whether there is a change in kidney function caused by hyperfiltration. We further performed a sex-stratified analysis, producing two models for male and female separately, using all the aforementioned variables apart from sex. All continuous variables, including the S2S distances, were standardised prior to being included in the regression models.

### Statistical Shape Analysis

Statistical shape analysis (SSA) is a technique defined by the variation of the size and shape (represented using a surface mesh) across participants. Principal component analysis (PCA) is a widely explored dimensionality reduction approach applied in SSA for constructing 3D statistical shape models [29, 30]. The principal directions of variation, also called modes of variation, could be represented by eigenvectors calculated from PCA. This technique finds a new coordinate system that describes the input data so that the greatest variance of the data lies on the first coordinate (the first principal component mode), the second-largest variance of the data that is orthogonal to the first component mode on the second coordinate, and so on. In this study, we performed PCA on the full cohort (N=38,868) to explore whether these coefficients for the principal components known as shape parameters can characterise variations in kidney shape across the population.

### Survival Analysis

Cox proportional hazards models were used to assess the risk of disease outcomes that occurred after the imaging visit. These models were adjusted for relevant covariates to account for potential confounding factors. To reduce the dimensionality of the S2S distances, we performed SSA by computing the PCA within each disease cohort and extracted the principal component (PC) scores.

To determine the unique contribution of each kidney measurement and ensure robust statistical significance of associations, we employed separate models for each kidney (left and right). This approach allowed us to account for potential confounding and effectively capture the specific associations between the kidney measures and disease outcomes. In these separate models, we adjusted for a comprehensive set of covariates including age, sex, ethnicity, body mass index, waist-to-hip ratio, alcohol intake frequency, smoking status, ibuprofen medication, sodium in urine, urea, kidney volume and the first four principal component scores of the S2S distances.

Disease outcomes and dates of the first occurrence of CKD, T2D, and hypertension were defined based on a combination of hospital records, primary care records, self-report, and death records (see Disease Definitions in supporting information and supplementary Table S1). Time-to-event was censored at the first event for each outcome, death, or last recorded follow-up (10th of February 2022), with a median of 3.7 years follow-up period since the imaging visit. Participants with an event recorded prior to the imaging visit were excluded.

Model summaries are reported as hazard ratios with 95% confidence intervals (CI). To control for multiple testing, the FDR procedure was estimated from the p-values, and a threshold of FDR < 0.05 determined significance.

## Results

### Study Population Characteristics

Of the cohort of 38,868 participants 96.9% were white and 48.3% male, with an age range between 44 and 82 years, and a mean BMI 26.5 ± 4.3 kg/m^2^ (supplementary Table S3). We identified 1,134 participants with CKD (581F/553M), age 69 ± 6.9 years and a BMI 27.8 ± 4.7 kg/m^2^, 2,054 participants with T2D from which 66% were male, age 66.6 ± 7.3 years and BMI 29.7 ± 5.2 kg/m^2^ and 14,113 participants with hypertension of which 58% male; age 66.8 ± 7.2 years; BMI 28 ± 4.6 kg/m^2^.

From all participants with CKD, defined by an eGFR below 60 ml/min/1.73 m², codes for chronic kidney disease, kidney dialysis, and kidney transplantation operation codes (supplementary Table S1), 466 (41.1%) participants had an eGFR below 60 ml/min/1.73 m² (53.8 ± 6.1, mean ± SD; 21.2 - 60 ml/min/1.73 m², range) and 793 participants were identified based on the codes for diagnosis. Hence, from the participants diagnosed with CKD (N=793), only 125 had eGFR below the aforementioned levels (supplementary Fig. S3). From the 14,113 participants with hypertension defined as self-reported of hypertensive medication, or a prior diagnosis of hypertension, or mean blood pressure ≥ 140/90 mmHg (see Disease Definitions in supporting information and supplementary Table S1), only 4,326 (30.7%) had mean blood pressure measurements ≥ 140/90 mmHg (supplementary Fig. S4).

### Associations with Anthropometric Characteristics and Disease

We proceeded to register the 200 healthy participant template on the full cohort (N=38,868), computing S2S distances between the template and surface of each individual kidney mesh, and performed MUR analysis to generate SPMs, adjusting for all relevant covariates. A summary of the model for the whole cohort, representing the standardised regression coefficients for each covariate and the significance areas on the kidney, is provided in Table 1, Table 2, and supplementary Fig. S5-S6. The SPMs that represent associations between S2S distances and the anthropometric measurements are shown in Fig. 1.

**Figure 1.**
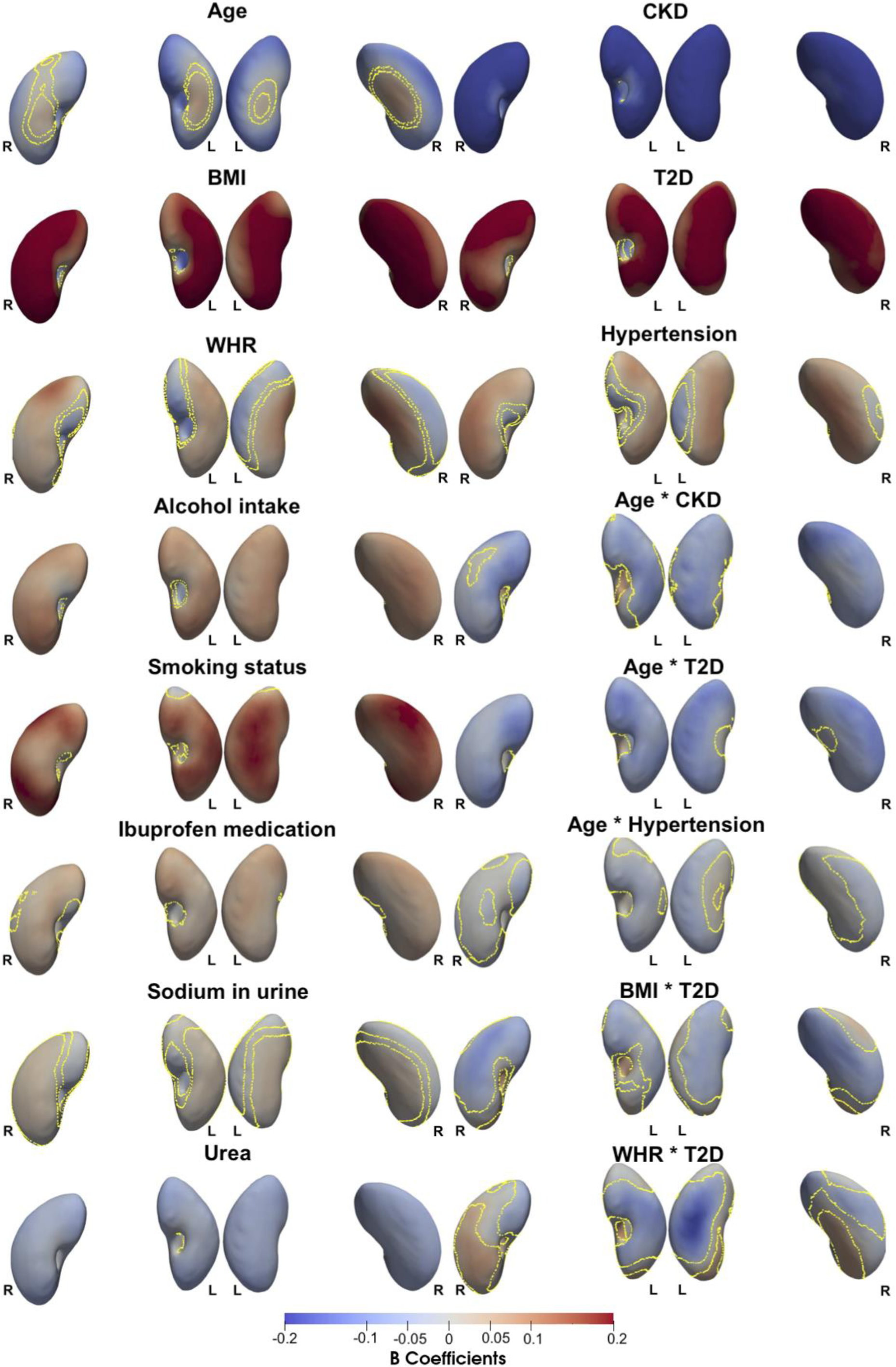
Three-dimensional statistical parametric maps (SPMs) of kidney morphology, projections are anterior and posterior views for both left (L) and right (R) kidneys in both anterior (left plots) and posterior (right plots) views. The SPMs show the local strength of association for each covariate in the model with S2S distances on the full cohort (N=38,868). Yellow contour lines indicate the boundary between statistically significant regions (p < 0.05) after correction for multiple testing, with positive associations in bright red and negative associations in bright blue. The standardised regression coefficients 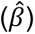 are shown with units in standard deviations for each covariate.

**Table 1.** Significance areas for covariates in the MUR model between the anthropometric covariates (N=38,868) in the model for the left kidney. The total area has been split into areas of positive, negative, and total associations. The standardised regression coefficients 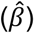 are presented as median (interquartile range - IQR) across all vertices of the left kidney surface and the significance areas as a percentage (%) of the vertices with statistically significant associations.

| Left Kidney | Standardised $\hat{\beta} < 0$ | | Standardised $\hat{\beta} > 0$ | | Total |
| --- | --- | --- | --- | --- | --- |
|  | Beta coefficients | Significance area | Beta coefficients | Significance area | Significance area |
| **Age** | -0.10 (0.07) | 89.84% | 0.03 (0.02) | 5.5% | 95.32% |
| **BMI** | -0.10 (0.07) | 14.79% | 0.20 (0.09) | 85% | 99.79% |
| **WHR** | -0.04 (0.04) | 37.44% | 0.04 (0.03) | 54.98% | 92.42% |
| **Alcohol intake frequency** | -0.06 (0.03) | 14.47% | 0.07 (0.02) | 84.43% | 98.9% |
| **Smoking status** | -0.05 (0.01) | 3.88% | 0.12 (0.05) | 84.36% | 88.24% |
| **Ibuprofen medication** | -0.04 (0.03) | 0.5% | 0.04 (0.02) | 84.27% | 84.77% |
| **Sodium in urine** | -0.02 (0.02) | 31.05% | 0.02 (0.01) | 48.81% | 79.86% |
| **Urea** | -0.06 (0.01) | 88.84% | 0.02 (0.02) | 6.6% | 95.43% |
| **CKD** | -0.33 (0.11) | 91.6% | 0.09 (0.04) | 3.74% | 95.34% |
| **T2D** | -0.06 (0.02) | 1.78% | 0.20 (0.08) | 85.5% | 87.28% |
| **Hypertension** | -0.03 (0.02) | 19.59% | 0.06 (0.04) | 67.44% | 87.03% |
| **Age \* CKD** | -0.06 (0.03) | 67.31% | 0.06 (0.02) | 9.54% | 76.85% |
| **Age \* T2D** | -0.06 (0.03) | 89.98% | 0.002 (0.002) | 0.07% | 90.05% |
| **Age \* Hypertension** | -0.03 (0.02) | 71.39% | 0.02 (0.003) | 1% | 72.4% |
| **BMI \* T2D** | -0.05 (0.02) | 59% | 0.05 (0.02) | 5.09% | 64.09% |
| **WHR \* T2D** | -0.07 (0.04) | 49.93% | 0.05 (0.02) | 8.97% | 58.9% |

**Table 2.** Significance areas for covariates in the MUR model between the anthropometric covariates (N=38,868) in the model for the right kidney. The total area has been split into areas of positive, negative, and total associations. The standardised regression coefficients 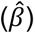 are presented as median (interquartile range - IQR) across all vertices of the left kidney surface and the significance areas as a percentage (%) of the vertices with statistically significant associations.

| **Right Kidney** | **Standardised $\hat{\beta} < 0$** |  | **Standardised $\hat{\beta} > 0$** |  | **Total** |
| --- | --- | --- | --- | --- | --- |
|  | Beta coefficients | Significance area | Beta coefficients | Significance area | Significance area |
| **Age** | -0.07 (0.05) | 78.32% | 0.03 (0.03) | 12.43% | 90.75% |
| **BMI** | -0.14 (0.03) | 12.46% | 0.23 (0.07) | 87.35% | 99.81% |
| **WHR** | -0.05 (0.07) | 27.26% | 0.04 (0.05) | 62.82% | 90.08% |
| **Alcohol intake frequency** | -0.05 (0.02) | 12.77% | 0.07 (0.02) | 85.77% | 98.54% |
| **Smoking status** | -0.06 (0.02) | 2.13% | 0.13 (0.05) | 92.17% | 94.3% |
| **Ibuprofen medication** | -0.01 (0.01) | 0.48% | 0.04 (0.01) | 83.21% | 83.68% |
| **Sodium in urine** | -0.02 (0.02) | 35.36% | 0.02 (0.01) | 48.78% | 71.61% |
| **Urea** | -0.06 (0.02) | 91.78% | 0.02 (0.02) | 4.91% | 96.69% |
| **CKD** | -0.30 (0.13) | 94.66% | 0.09 (0.03) | 2.54% | 97.29% |
| **T2D** | -0.06 (0.02) | 6.97% | 0.20 (0.05) | 87.76% | 94.73% |
| **Hypertension** | -0.04 (0.03) | 10.76% | 0.07 (0.05) | 79.76% | 90.51% |
| **Age \* CKD** | -0.06 (0.04) | 72.54% | 0.06 (0.02) | 3.74% | 76.28% |
| **Age \* T2D** | -0.06 (0.03) | 89.84% | - | - | 89.84% |
| **Age \* Hypertension** | -0.02 (0.01) | 40.97% | 0.01 (0.01) | 0.19% | 41.16% |
| **BMI \* T2D** | -0.07 (0.04) | 59.63% | 0.06 (0.03) | 14.23% | 73.86% |
| **WHR \* T2D** | -0.05 (0.02) | 22.28% | 0.05 (0.03) | 28.56% | 50.84% |

S2S distances were negatively associated with age, showing a median shape variation of −0.1 covering over 89.8% of the left kidney, and a median shape variation of −0.08 covering 78.3% of the right kidney, observed in the inferior and superior areas of both kidneys. There was also a positive association between age and S2S distances with a median shape variation of 0.03 covering only 5.5% of the left kidney and a median shape variation of 0.03 covering 12.4% of the right kidney, shown in the anterior and posterior areas of both kidneys. BMI and WHR had statistically significant positive associations with S2S distances, covering 85% and 55% of the left kidney surface and 87.3% and 62.8% of the right kidney, respectively. Alcohol intake frequency showed a statistically significant outward shape variation with a median of 0.07 in both kidneys, whereas smoking status showed a statistically significant outward shape variation with a median of 0.12 in the left kidney and 0.13 in the right kidney. Ibuprofen medication showed a median outward shape variation in S2S distances of 0.04 in both kidneys (significance area = 84.3% for the left kidney and 83.2% for the right kidney). We further examined the associations between S2S distances and biological markers including sodium in urine and urea. Sodium in urine showed a statistically significant positive association with S2S distances with a median shape variation of 0.02 (significance area = 48.8%) in both kidneys. Urea showed a median inward shape variation with S2S distances of −0.06 (significance area = 88.8%) in the left kidney and −0.06 (significance area = 91.8%) in the right kidney.

A diagnosis of CKD was associated with S2S distances with a median shape variation of −0.33 (significance area = 91.6%) in the left kidney and −0.30 (significance area = 94.7%) in the right kidney. T2D was positively associated with S2S distances, with a median shape variation of 0.20 in both kidneys covering a significant area of 85.5% of the left kidney and a significant area of 87.8% of the right kidney. Finally, hypertension showed median outward shape variation in S2S distances of 0.06 covering 67.4% of the left kidney and 0.07 covering 79.8% of the right kidney.

We undertook further analysis to determine whether there was an interaction between all clinical conditions and age-adjusted for all covariates in the model. We observed a stronger relationship between age and S2S distances with a median inward shape variation of −0.16 in CKD participants, compared with −0.10 in non-CKD participants in the left kidneys covering an area of 67.3% and median shape variation in S2S distances of −0.13 in CKD participants, covering an area of 72.5% compared with −0.07 in non-CKD participants in the right kidney (Table 2,3 and Fig. 1). The association between age and S2S distances in participants with CKD and without a clinical condition are directly compared in Fig. 2i and 2ii. Participants with T2D (Fig. 2iii) display a stronger relationship with S2S distances with a median inward shape variation of −0.16 in 90% of the left kidney and a median inward shape variation of −0.12 in 89.8% of the right kidney with increasing age. Participants with hypertension (Fig. 3iv) showed a similar relationship with S2S distances with increasing age with a median inward shape variation of −0.13 covering an area of 71.4% in the left kidney and with a median inward variation of −0.09 in 41% of the right kidney.

**Figure 2.**
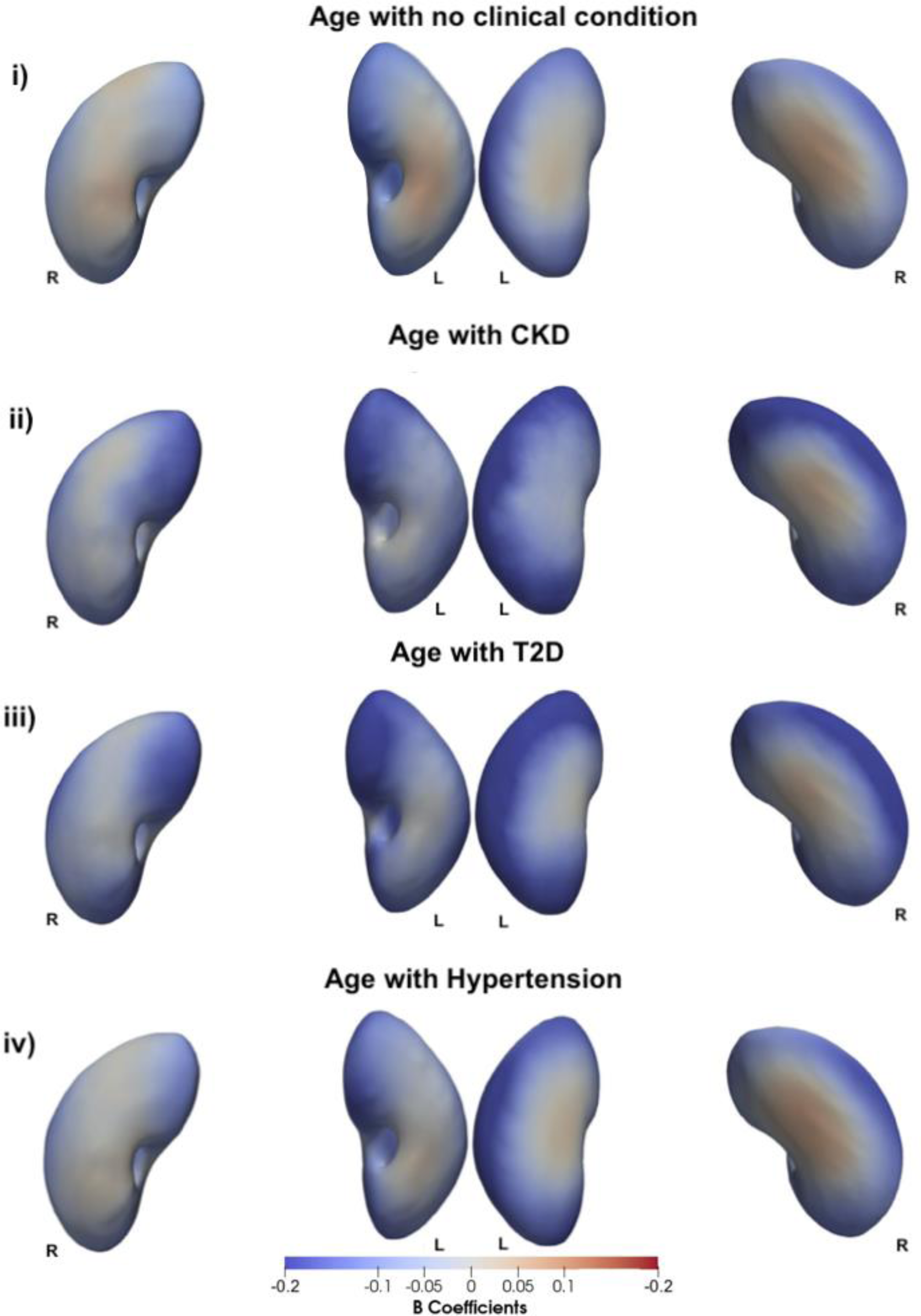
Three-dimensional statistical parametric maps (SPMs) of kidney morphology, projections are anterior (left plots) and posterior (right plots) views for both left (L) and right (R) kidneys. The SPMs show the local rate of change as a function of age for S2S distances in participants **i)** without CKD, T2D, or hypertension, **ii)** with CKD, **iii)** with T2D, and **iv)** with hypertension in the full cohort (N=38,868). Positive associations are in red and negative associations are in blue. Regression coefficients 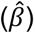 are shown with units in standard deviations.

**Figure 3.**
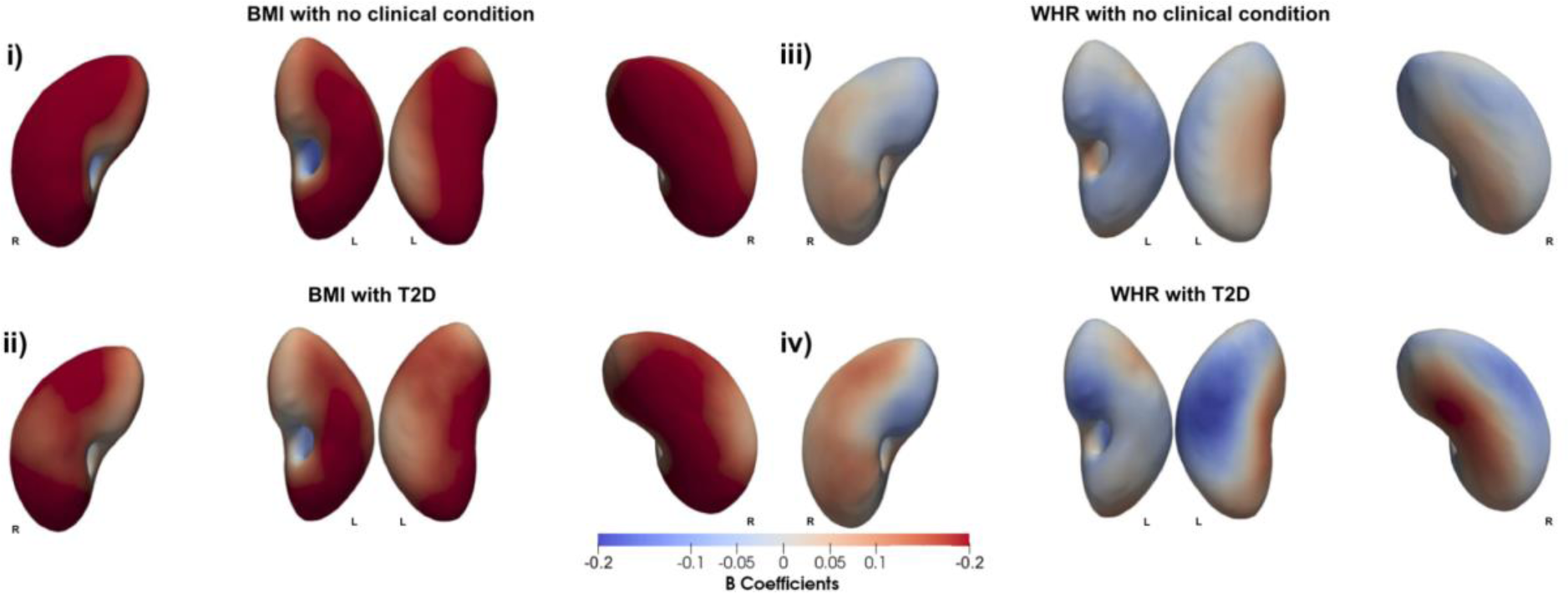
Three-dimensional statistical parametric maps (SPMs) of kidney morphology, projections are anterior (left plots) and posterior (right plots) views for both left (L) and right (R) kidneys. **i)** The SPMs show the local rate of change as a function of BMI for S2S distances in participants without T2D and **ii)** with T2D, **iii)** SPMs show the local rate of change as a function of WHR for S2S distances in participants without T2D and **iv)** with T2D on the full cohort (N=38,868). Positive associations are in red and negative associations in blue. Regression coefficients 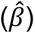 are shown with units in standard deviations.

We further investigated whether there was an interaction between T2D and BMI as well as WHR adjusted for all covariates in the model (Table 1, 2, and Fig. 1). BMI in participants with T2D showed a stronger relationship with S2S distances with a median outward shape variation 0.05 covering only a small significance area of only 5.1% of the left kidney and a median outward shape variation of 0.06 covering an area of 14.2% of the right kidney in addition to the median outward shape variation of 0.20 in the left kidney (significance area = 85%) and 0.23 in the right kidney (significance area = 87.4%) for the main effect of BMI. However, we observed a stronger relationship in S2S distances with a median inward shape variation of −0.05 over an area of 59% in the left kidney and a median inward shape variation of −0.07 covering an area of 59.6% in the right kidney for the main effect of BMI (Fig. 3i, 3ii).

From the interaction between WHR and T2D (Tables 1 and 2, and Fig. 1), we observed a stronger relationship with S2S distances with a median outward shape variation of 0.05 in the left kidney (significance area = 9%) and 0.05 in the right kidney (significance area = 28.6%), in addition to the median shape variation of 0.04 in the left kidney (significance area = 55%) and 0.04 in the right kidney (significance area = 62.8%) for the main effect of WHR. However, we also found a stronger relationship in S2S distances with a median inward shape variation of −0.07 (significance area = 50%) in addition to the median shape variation of −0.04 (significance area = 37.4%) in the left kidney and −0.05 (significance area = 22.3%) in addition to the median shape variation of −0.05 (significance area = 27.3%) in the right kidney for the main effect of WHR (Fig. 3iii, 3iv).

### Associations by Sex

We performed MUR analysis to explore the associations between kidney shape and anthropometric and disease traits separated by sex, adjusting for all relevant covariates excluding sex. A summary of the model for the whole cohort, representing the standardised regression coefficients and the significance areas on the kidney is provided in supplementary Table S4, and Table S5, and the histograms showing the statistically significant standardised regression coefficients is provided in supplementary Fig. S7-S8. The SPMs that represent associations between S2S distances and the anthropometric measurements with units in standard deviations for each covariate are shown in Fig. S9 for male (N=18,855) and Fig. S10 for female participants (N=20,013).

We observed similar associations with the model including the full cohort for both sexes. However, we noticed that the WHR in both the left and right kidneys showed more statistically significant negative associations with S2S distances in male participants (median inward shape variation= −0.05 in both kidneys; significance area = 71.4% in the left kidney; 45.5% in the right kidney) and more statistically significant positive associations in female participants (median outward shape variation = 0.05, significance area = 74.9% in the left kidney; median outward shape variation = 0.04, 66.2% in the right kidney) (supplementary Table S4, Table S5).

We also observed the interaction between age and all disease outcomes, BMI and T2D as well as WHR and T2D for male and female participants, separately. Female participants with CKD showed a strong relationship with S2S distances with a median inward shape variation of −0.09 in the left kidney (significance area = 29.8%) and −0.08 in the right kidney (significance area = 65.8%), in addition to the median shape variation of −0.10 (significance area = 85.3%) in the left kidney and −0.08 (significance area = 74.1%) in the right kidney for the main effect of age. While, male participants with CKD showed a strong relationship with S2S distances with a median inward shape variation of −0.10 in the left kidney (significance area = 44.8%) and −0.11 in the right kidney (significance area = 18.5%), in addition to the median shape variation of −0.10 (significance area = 91.1%) in the left kidney and for −0.08 (significance area = 79.4%) in the left kidney the main effect of age. We further found that both male and female participants with T2D showed a strong relationship with S2S distances showing a greater significance area in male participants with a median inward shape variation of −0.08 (significance area = 82%) in the left kidney and −0.07 in the right kidney (significance area = 85.6%), for the main effect of age. Furthermore, male participants with hypertension showed a stronger relationship with S2S distances for the main effect of age, with a median inward shape variation of −0.03 in both kidneys (significance area = 40.3% in the left kidney and 49.6% in the right kidney).

Interactions between BMI and T2D showed a strong relationship with S2S distances in both sexes although, a greater significance area was observed in men with a median inward shape variation of −0.08 for both kidneys covering a significance area of 67% of the left kidney and 80.5% of the right kidney. Finally, male participants with T2D showed a strong relationship with S2S distances for the main effect of WHR, with a median outward shape variation of 0.07 in the left kidney (significance area = 15.4%) and a median outward shape variation of 0.08 in the right kidney (significance area = 22.6%), while female participants with T2D showed a median inward shape variation of −0.08 in the left kidney (significance area = 28.9%) for the main effect of WHR.

### Statistical Shape Analysis

To visualise shape variation across a population, we computed the PCA of the coordinate system including the S2S distances from the full cohort (N=38,868). The first four modes of kidney size and shape variation from the PCA are presented as −3 SD, mean and +3 SD (Fig. 4i and supplementary video S1).

**Figure 4.**
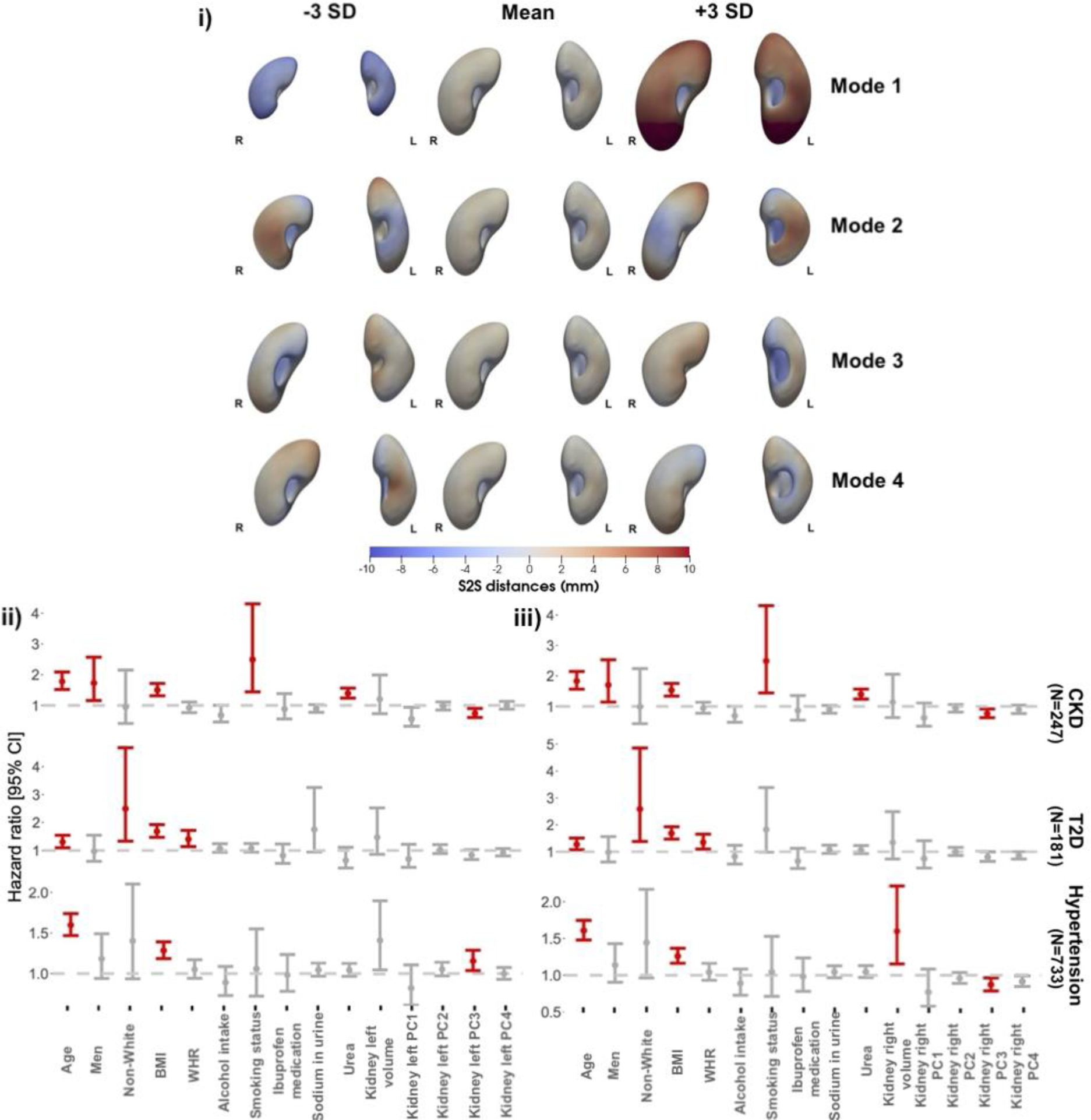
**i)** The first four modes of shape variation for the kidneys of the full cohort (N=38,868). The mean shape and the shape at the +/- three SD are displayed for each mode showing the S2S distance variation in mm. The kidney shape variations are shown in the anterior views for both left (L) and right (R) kidneys. **ii)** Hazard ratios and 95% CIs for the three outcomes: CKD, T2D, and hypertension for the left kidney. **iii)** Hazard ratios for the three disease outcomes for the right kidney adjusted for age, sex, ethnicity, body mass index, waist-to-hip ratio, sodium in urine, urea, alcohol intake frequency, smoking status, ibuprofen medication uptake, kidney volume and the first 4 principal component scores for the S2S distances. Significant associations for p < 0.05 are shown in red and non-significant associations in grey.

The principal components are mathematically derived, so they do not necessarily correspond to physical features but they can be broadly interpreted by visualising extreme values along each axis (+/- 3 SD). The percentage of shape variation explained by the first ten modes of PCA for both left and right kidneys on the full cohort is presented in supplementary Fig. S11. The first principal component accounted for 39.5% of the shape variation for the left kidney and 41.2% for the right kidney and appeared to correspond best to the size of the kidney. The second principal component accounting for 7.5% of the shape variation for the left kidney and 8% for the right kidney, appeared to correspond to the angle of the superior end and the straightness of the kidney whereas the third principal component accounting for 6.5% of the shape variation for the left kidney and 5.6% for the right kidney, is visually associated with the length and width of the kidney. The fourth principal component appeared to be associated with the length and thickness (accounting for 3.8% of the shape variation for the left kidney and 4.4% for the right kidney). The remaining principal components are difficult to visually interpret.

### Survival Analysis

We assessed whether the S2S distances had predictive value for the three disease outcomes diagnosed after the imaging visit. Here we identified 247 participants with CKD of which 59% were male, aged between 68.6 ± 6.5 years (mean ± SD) and a BMI between 16.4 and 48.9 kg/m^2^, 181 participants with T2D of which 61% were male, aged 65.4 ± 7.6 years with a BMI between 19.5 and 47.9 kg/m^2^ and finally, 733 participants with hypertension (56.2% male; aged 65.8 ± 7.1 years; BMI from 17.4 to 53 kg/m^2^). We created a model for each disease outcome adjusting for age, sex, ethnicity, body mass index, waist-to-hip ratio, alcohol intake frequency, smoking status, ibuprofen medication, sodium in urine, urea, kidney volume, and the first 4 PC scores for the S2S distances (accounting for over 60% of the variation in S2S distances in all disease outcomes and for both kidneys), separately for each kidney (Fig. 4ii for the left kidney; Fig. 4iii for the right kidney).

We found that the third PC scores of the S2S distances were risk factors for CKD diagnosis in both the left kidney (0.74 [0.61-0.90]) and the right kidney (0.76 [0.63-0.92]). No PC scores of the S2S distances in both left and right kidneys were associated with T2D. This may be attributed to the relatively short follow-up period from the imaging visit (median 3.7 years, IQR 2.3 years) as well as the small number of incidences of T2D diagnosis (N=181), resulting in reduced statistical power to detect significant associations.

We further found that increased right kidney volume was a risk factor for hypertension (1.60 [1.15-2.22]). The third PC score of the S2S distances for the left kidney was significantly positively associated with risk of hypertension (1.16 [1.04-1.29]) however, PC3 score of the S2S distances for the right kidney was significantly negatively associated with risk of hypertension (0.87 [0.79-0.96]).

## Discussion

In this study, we mapped local shape variations across the kidneys and determined how these variations were associated with anthropometric and disease traits. To achieve this we constructed surface meshes from kidney segmentations of 38,868 participants from the UK Biobank. Previous studies using similar SPM techniques have suggested that this is a useful technique in neuroimaging [14] and cardiac imaging [16], for mapping the associations between phenotypic and genetic variation in specific anatomical regions.

Measurements of the kidneys have been extensively explored using a variety of approaches from computed tomography angiography (CTA) [31], ultrasound [6], computed tomography (CT), and MRI [32], with assessments including measurements of the whole organ as well as sub-structures including kidney cortex (the outer layer), medulla (the inner layer) and parenchyma (the tissue that forms the bulk of the kidneys composed of nephrons), and typically include measurement of size, length [33, 34], volume [3] and cortical thickness [35]. While these methods enhance our understanding of the kidneys in population-level studies, they may not be able to capture specific morphological and regional variations that occur in the kidneys in response to specific conditions. The SPM method implemented in this study demonstrates significant regional variations in each participant’s kidney shape relative to an average kidney shape such that an outward or inward variation would add or subtract from the surface and illustrates their associations with anthropometric variables and disease outcomes. We believe that these novel phenotypic variables can be useful in longitudinal population studies, as well as in determining trajectories of progression in clinical conditions involving the kidneys, including CKD.

Our study provides an insight into the morphometric variations of both left and right kidney surface meshes and quantifies the anatomical relationships with age, sex, body composition, lifestyle, and disease including CKD, T2D, and hypertension. We demonstrate that age was significantly associated with a decrease in S2S distances observed in the inferior and superior areas of both kidneys and was positively associated with S2S distances in the anterior and posterior areas of the kidneys. Previous studies investigating the kidney sub-structures of potential kidney donors found that cortical volume decreases with age and increases in medullary volume, with a slight difference in females [3]. Other studies investigating the parenchymal and total kidney volume of healthy individuals, found that kidney volume and length increased up to middle age followed by a continuous decrease in men and slowly decreased in women, suggesting some progressive loss of functional nephrons [36]. In our analysis, we found a similar decline in S2S distances with age for both sexes. However, future work will help to shed light on the shape variations from the kidney sub-structures such as the cortex and medulla.

We further found statistically significant positive associations between S2S distance and both BMI and WHR. Previous reviews of the associations between kidney measurements and BMI for different age, sex, and ethnicity groups, reported that the most significant factor related to kidney volume is BMI [37]. They also show that in most studies BMI showed a positive correlation with kidney size, length, or volume. Lifestyle factors including alcohol intake frequency and smoking were positively associated with S2S distances, showing median outward shape variations. Indeed, increased kidney volume has previously been associated with current smoking [3]. Furthermore, it has been shown that both smoking and heavy drinking are associated with kidney dysfunction and can contribute to the risk of glomerular hyperfiltration, CKD progression, and dementia [38, 39].

We found that the presence of CKD was associated with reduced S2S in both kidneys. From previous sonographic measurements of the kidneys, it has been shown that a reduction in kidney volume can be used as an indicator of CKD [40], enabling the use of kidney size for the prediction of kidney function in healthy populations as well as patients with CKD [41, 42]. We further identified significant outward shape variations with the presence of T2D and hypertension which was consistent in both kidneys. It is known from previous studies that patients with non-insulin dependent diabetes mellitus develop enlarged kidneys with hyperfiltration [43]. Further studies investigating the associations of hyperfiltration and hypofiltration with prediabetes and prehypertension show both diabetes and hypertension associate with hyperfiltration [44] however, another study showing independent associations between kidney volume and diastolic blood pressure did not find a significant association [3].

Interaction models within disease groups showed a stronger relationship between age and S2S distances in both kidneys with inward shape variation for all disease outcomes. More specifically our findings demonstrate that the added effect of age and CKD may further explain the inward shape variations in the kidneys’ S2S distances. This could be explained by the loss of nephrons, which lead to the progressive loss of kidney function as CKD progresses [45]. BMI and WHR also showed a stronger relationship with shape variations in participants with T2D. We also considered whether there are similar interactions for both sexes separately. When we considered the interaction between age and CKD, we found a stronger relationship with S2S distances with inward shape variation in both male and female participants. Both male and female participants with T2D showed a stronger relationship with S2S distances with inward shape variation for the main effect of age. Interactions between BMI and T2D showed a stronger relationship with S2S distances for both sexes although a greater significance area was observed in men. It is worth noting that due to the clinical heterogeneity of our current clinical cohort (CKD, T2D and hypertension), in terms of time of diagnosis and medication, as well as the possibility of collider bias or reverse confounding, it is impossible to identify causal mechanisms for the observed variation in S2S distances.

We further explored variations in kidney morphometry and demonstrated that the first four principal components of kidney shape were associated with size, angle, straightness, width, length and thickness of the kidneys. These observations are similar to the findings in [11], reporting that the kidney shape variations are heavily biased by organ size, angle, straightness, length-to-width ratio and length-to-thickness ratio. The rationale for this analysis was to capture SSA-based shape features of the kidneys that would be independent from current existing clinically available predictors such as kidney volume. However, future work is needed to condense the entire coordinate matrix or deformation conventional matrix into most distinct modes to categorise population variations, which could be used in genetic association studies and enhance our understanding of kidney related diseases [46, 47].

We also investigated the risk of future disease outcomes adjusted for relevant anthropometric variables, kidney volumes as well as kidney S2S distances. Since S2S distances include high-dimensional data of vertices on the mesh, we applied PCA across all participants at each disease cohort and included the first four PC scores of the S2S distances, as they provide interpretable components. Our findings revealed contrasting associations between the SSA-based S2S variations corresponding to the length and width in the left and right kidneys with risk of hypertension. Specifically, we observed a significant positive association in the left kidney but a negative association in the right kidney. This difference may relate to the anatomical variations that predispose to atheroma in the renal artery which may then contribute to the associations with hypertension. This discrepancy may be also attributed to the significant anatomical differences in kidney length and width between the left and right sides. A previous study utilising sonographic measurements of renal size have also reported such disparities [48]. They also suggest that these differences may be attributed to the greater spatial capacity in the left kidney and a shorter length of the left renal artery compared to the right renal artery, resulting in increased blood flow and potentially an increase in kidney volume. We further demonstrated that variations corresponding to the length and width in the left and right kidney were associated with incidents of CKD. Previous studies measuring kidney size by ultrasonography, reported that kidney length and volume were correlated with eGFR levels in the elderly, however they report that kidney length had lower specificity in predicting kidney dysfunction [42]. Other studies investigating the accuracy of sonographic kidney measurements to detect kidney impairment and histological change, reported that kidney length to height ratio weighted for kidney echogenicity was able to detect kidney dysfunction [49]. Although these studies report the usefulness of the sonographic kidney measurements in detecting loss of kidney function, our study highlights the importance of the SSA-based shape features, in predicting the risk of future disease outcomes. More longitudinal measurements and relevant outcome data will be required to evaluate whether the S2S variations alone are a useful measure in predicting risk of kidney disease.

Our study has limitations. The UK Biobank is a large cross-sectional study that is subject to selection bias with a “healthier” cohort than the wider UK population, who are predominantly of European ancestry, excludes younger participants and potentially more severe cases [50, 51]. However, it has been shown that risk factor associations are likely to be generalisable [52]. Another potential limitation of this study is that to ensure sufficient numbers of participants in the CKD group, we included both participants based on CKD diagnosis codes reported at the imaging visit and participants based on eGFR levels taken at the initial assessment visit which precedes the imaging visit of 9 years. Also, although albumin/creatinine ratio (ACR) is widely used to assess kidney function and diagnose diseases such as CKD, cardiovascular disease and diabetes [53, 54], this parameter was not used in this study as urine albumin was only available for 28% of the UK Biobank imaging cohort [55, 56]. Furthermore, this study has only a 3.7 years follow-up period since the imaging visit, which may limit the power of time-to-event studies. Additional longitudinal measurements will need to be required to assess age-related changes in disease cohorts.

## Conclusions

Our findings highlight the potential scientific and clinical significance of shape analysis techniques. We found statistically significant associations between S2S distances and disease outcomes including CKD, T2D and hypertension. We also identified significant associations between the SSA-based shape features corresponding to the size, length and width, and future events of disease outcomes. Therefore, the application of SPM and SSA-based shape analysis is feasible for improving our understanding of the variations in kidney shape associated with disease outcomes and their prediction. These techniques will benefit future research in population-based cohort studies, in identifying associations between physiological, genetic and environmental effects on structure and function of the kidneys as well as the kidney sub-structures such as cortex and medulla.

## Data Availability

The data that support the findings of this study are available from the UK Biobank (https://www.ukbiobank.ac.uk), but restrictions apply to the availability of these data, which were used under license for the current study, and so are not publicly available. Data are however returned by us to the UK Biobank where they will be fully available on request.

https://www.ukbiobank.ac.uk

## Abbreviations

CKD: chronic kidney disease
T2D: type 2 diabetes
BMI: body mass index
WHR: waist-to-hip ratio
eGFR: estimated glomerular filtration rate
MUR: mass univariate regression
TFCE: threshold-free cluster enhancement
SPMs: statistical parametric maps
SSA: Statistical Shape Analysis
PCA: principal component analysis
S2S: Surface-to-surface.

## Declarations

### Ethics approval and consent to participate

The data resources used in this study have approval from ethics committees. Full anonymised images and participants metadata from the UK Biobank cohort was obtained through UK Biobank Access Application number 44584. The UK Biobank has approval from the North West Multi-Centre Research Ethics Committee (REC reference: 11/NW/0382), and obtained written informed consent from all participants prior to the study. All methods were performed in accordance with the relevant guidelines and regulations as presented by the relevant authorities, including the Declaration of Helsinki https://www.ukbiobank.ac.uk/learn-more-about-uk-biobank/about-us/ethics.

### Consent for publication

Not applicable.

### Author Contributions

J.D.B., E.L.T., M.T. and M.C. conceived the study. R.L., J.D.B., B.W., E.L.T., and M.T. designed the study. M.T. and B.W. implemented the methods and performed the data analysis. M.T. defined the disease and physiological condition categories. M.T. performed the image and statistical analysis. R.L., E.L.T., B.W., M.T., J.D.B., E.P.S., R.S. and N.B. drafted the manuscript. All authors read and approved the manuscript.

## Acknowledgements

We thank Alex Chekholko for providing technical support that expedited our work. This research has been conducted using the UK Biobank Resource under Application Number 44584.

