## Supplementary_Material for "Kidney Shape Statistical Analysis: Associations with Disease and Anthropometric Factors"

Disease Definitions

Disease categories of interest, including CKD, T2D and hypertension were selected based on their influence on body composition and organ health, as well as their frequency within the UK Biobank. They were defined at the time and before the imaging visit. A summary of the codes corresponding to the considered disease traits are provided in Supplementary Table S1.

For the definition of CKD, we calculated eGFR using the CKD-EPI equation (see methods section) and baseline cohort data. In a previous study [[1]](https://paperpile.com/c/KvRrXW/HZdm), mild to moderate CKD was defined by an eGFR between 15 and 60 ml/min/1.73 m² or by the International Classification of Disease (ICD) codes N18.1-N18.5. We define CKD by eGFR below 60 ml/min/1.73 m², and the ICD10 and self-reported codes for chronic kidney disease, kidney dialysis and kidney transplantation operation codes (<https://www.datadictionary.nhs.uk/Covid19PRA/Chronic_Kidney.html>). The codes for T2D were selected based on the ICD10 and self-reported codes for type-2 diabetes [[2]](https://paperpile.com/c/KvRrXW/ZSqO). Hypertension was defined as self-report of hypertensive medication use, or a prior diagnosis of hypertension, or mean blood pressure ≥ 140/90 mmHg [[3]](https://paperpile.com/c/KvRrXW/tJJq). The codes for hypertension diagnosis were selected based on the ICD10 and the self-reported codes for hypertension. Hypertensive medication codes were selected from the medication fields, reporting as regularly taking blood pressure medication.

CKD, T2D and hypertension diagnosed or reported after the imaging visit was defined based on ICD10 codes from the various sources, the date of first diagnosis made available as “first occurrence” data (Category 1712) and based on self-reported codes described in Supplementary Table S1, defined after the imaging visit.

Supplementary Tables

| **Trait** | **ICD-10** | **OPCS-4** | **Self-Reported Code (20002)** | **Operation Code**  **(20004)** | **UK Biobank Field** |
| --- | --- | --- | --- | --- | --- |
| **CKD** | N18 | M01, M011-M015,  M018-M019, X401-X406, X408-X409 | 1193 | 1195, 1580, 1581, 1582 | - |
| **T2D** | E11 | - | 1220, 1223 | - | - |
| **Hypertension** | I10-I13, I15, O10 | - | 1065, 1072 | - | 6153, 6177 |

**Table S1.** Summary of the codes used to define disease.

|  | **Full Cohort**  (N = 200) | **Male**  (N = 100) | **Female**  (N = 100) |
| --- | --- | --- | --- |
| **Age** $(yrs.)$ | 53.76 ± 3.71  (46, 62) | 55.13 ± 3.56  (48, 62) | 52.39 ± 3.35  (46, 62) |
| **Weight** $(kg)$ | 65.68 ± 8.17  (46.2, 90.1) | 70.85 ± 6.65  (57.5, 90.1) | 60.50 ± 6  (46.2, 76.8) |
| **Height** $(m)$ | 1.72 ± 0.09  (1.53, 1.93) | 1.78 ± 0.07  (1.61, 1.93) | 1.66 ± 0.06  (1.51, 1.80) |
| **BMI** ${(kg/m}^{2})$ | 22.17 ± 1.56  (18.5, 24.90) | 22.41 ± 1.50  (18.5, 24.90) | 21.93 ± 1.59  (18.6, 24.77) |
| **Waist circumference** $(cm)$ | 76.84 ± 7.64  (60, 93) | 81.75 ± 5.79  (68, 93) | 71.94 ± 5.92  (60, 91) |
| **Hip circumference** $(cm)$ | 93.71 ± 4.7  (82, 111) | 94.17 ± 4.74  (85, 111) | 93.24 ± 4.63  (82, 103) |
| **WHR** | 0.82 ± 0.07  (0.67, 0.98) | 0.87 ± 0.05  (0.74, 0.98) | 0.77 ± 0.05  (0.67, 0.93) |
| **SBP** $(mmHg)$ | 112.04 ± 6.21  (92.5, 120) | 113.45 ± 5.15  (95, 120) | 110.63 ± 6.86  (92.5, 120) |
| **DBP** $(mmHg)$ | 69.28 ± 4.76  (60, 80) | 69.68 ± 4.67  (60, 80) | 68.88 ± 4.84  (60.5, 80) |
| **Sodium in urine** $(mmol/L)$ | 63.6 ± 38.68  (10.5, 193.1) | 72.3 ± 42.13  (13.3, 193.1) | 54.9 ± 32.86  (10.5, 154.6) |
| **Urea** $(mmol/L)$ | 4.88 ± 1.22  (1.46, 9.45) | 5.19 ± 1.38  (1.46, 9.45) | 4.58 ± 0.96  (2.71, 6.88) |
| **eGFR** $(ml/min/1.73 m^{2})$ | 99.76 ± 11.00  (66.31, 127.95) | 102.02 ± 9.85  (77.56, 127.95) | 97.18 ± 12.21  (66.31, 119.14) |
| **Alcohol intake frequency** $(n)$ | 8 | 2 | 6 |
| **Smoker** $(n)$ | 6 | 3 | 3 |
| **Ibuprofen medication** $(n)$ | 36 | 9 | 27 |
| **Left kidney volume** $(ml$) | 135.31 ± 23.2  (71.48, 202.02) | 147.12 ± 19.63  (98.11, 202.02) | 123.5 ± 20.36  (71.48, 181.98) |
| **Right kidney volume** $(ml$) | 129.19 ± 23.03  (53.00, 189.50) | 141.36 ± 19.99  (97.81, 189.50) | 117.02 ± 19.16  (53.00, 162.13) |

**Table S2.** Summary statistics (mean ± standard deviation, minimum and maximum values) for continuous variables and counts for discrete variables of the 200-participant gender-balanced cohort for the template construction. BMI: body mass index, WHR: waist-to-hip ratio, SBP: Systolic blood pressure, DBP: Diastolic blood pressure, eGFR: estimated glomerular filtration rate.

|  | **Full Cohort** | **Male**  (N = 18,855) | **Female**  (N = 20,013) |
| --- | --- | --- | --- |
| **Caucasian** $(n)$ | 37,632 | 18,226 | 19,406 |
| **Age** $(yrs.)$ | 64.14 ± 7.71  (44, 82) | 64.84 ± 7.8  (44, 82) | 63.49 ± 7.57  (45, 82) |
| **Weight** $(kg)$ | 76.06 ± 15.04  (34.3, 169.2) | 83.65 ± 13.21  (46.5, 169.2) | 68.91 ± 13.01  (33.3, 169.2) |
| **Height** $(m)$ | 1.69 ± 0.09  (1.34, 2.04) | 1.76 ± 0.07  (1.5, 2.04) | 1.63 ± 0.06  (1.34, 1.95) |
| **BMI** ${(kg/m}^{2})$ | 26.48 ± 4.34  (14.08, 62.04) | 26.96 ± 3.84  (16.48, 53.91) | 26.03 ± 4.72  (14.08, 62.04) |
| **Waist circumference** $(cm)$ | 88.36 ± 12.65  (53, 184) | 94.31 ± 10.63  (63, 184) | 82.75 ± 11.80  (53, 144) |
| **Hip circumference** $(cm)$ | 100.74 ± 8.69  (71, 176) | 100.68 ± 7.30  (76.2, 176) | 100.79 ± 9.83  (71, 157) |
| **WHR** | 0.88 ± 0.09  (0.53, 1.47) | 0.94 ± 0.06  (0.53, 1.47) | 0.82 ± 0.07  (0.60, 1.22) |
| **SBP** $(mmHg)$ | 138.97 ± 18.60  (76.5, 240.5) | 142.11 ± 17.4  (76.5, 240.5) | 135.99 ± 19.21  (77.5, 227.5) |
| **DBP** $(mmHg)$ | 78.75 ± 10.05  (37.5, 129) | 80.58 ± 9.86  (46.5, 129) | 77.01 ± 9.92  (37.5, 121.5) |
| **Sodium in urine** $(mmol/L)$ | 73.91 ± 41.13  (10, 316.7) | 84.25 ± 42.36  (10, 306.9) | 64.17 ± 37.42  (10, 316.7) |
| **Urea** $(mmol/L)$ | 5.32 ± 1.21  (1.5, 19.38) | 5.54 ± 1.22  (1.81, 19.38) | 5.12 ± 1.18  (1.5, 19.2) |
| **eGFR** $(ml/min/1.73 m^{2})$ | 91.66 ± 12.18  (21.22, 133.80) | 91.10 ± 11.97  24.18, 131.91) | 92.19 ± 12.35  (21.22, 133.80) |
| **Alcohol intake frequency** $(n)$ | 6,659 | 3,932 | 2,727 |
| **Smoker** $(n)$ | 1,326 | 759 | 567 |
| **Ibuprofen medication** $(n)$ | 4,164 | 1,657 | 2,507 |
| **Left kidney volume** $(ml$) | 135.07 ± 30.33  (30.62, 357.24) | 148.61 ± 30.36  (30.62, 357.24) | 122.32 ± 24.16  (31.10, 271.51) |
| **Right kidney volume** $(ml$) | 135.44 ± 22.22  (30.70, 378.09) | 149.66 ± 28.06  (30.70, 378.09) | 122.05 ± 23.40  (31.67, 335.41) |

**Table S3.** Summary statistics (mean ± standard deviation, minimum and maximum values) for continuous variables and counts for discrete variables in the full cohort (N=38,868). BMI: body mass index, WHR: waist-to-hip ratio, SBP: Systolic blood pressure, DBP: Diastolic blood pressure, eGFR: estimated glomerular filtration rate.

| **Left Kidney** | **Male** | | **Female** | |
| --- | --- | --- | --- | --- |
|  | $\hat{\beta}<0$ | $\hat{\beta}>0$ | $\hat{\beta}<0$ | $\hat{\beta}>0$ |
|  | Median Beta coefficients (IQR, Significance area (%)) | | | |
| **Age** | -0.10 (0.08, 91.1%) | 0.03 (0.01, 2.03%) | -0.10 (0.06, 85.3%) | 0.04 (0.02, 8.97%) |
| **BMI** | -0.12 (0.09, 13.6%) | 0.21 (0.13, 86.1%) | -0.10 (0.08, 15.1%) | 0.22 (0.12, 84.5%) |
| **WHR** | -0.05 (0.05, 71.4%) | 0.04 (0.03, 9.27%) | -0.04 (0.04, 14.5%) | 0.05 (0.04, 74.9%) |
| **Alcohol intake frequency** | -0.05 (0.03, 12.3%) | 0.06 (0.03, 84.3%) | -0.08 (0.03, 14.8%) | 0.11 (0.04, 82.9%) |
| **Smoking status** | -0.07 (0.01, 0.39%) | 0.13 (0.07, 86.1%) | -0.09 (0.03, 3.29%) | 0.15 (0.09, 72.9%) |
| **Ibuprofen medication** | -0.05 (0.02, 0.64%) | 0.07 (0.02, 83.9%) | - | 0.04 (0.02, 46.4%) |
| **Sodium**  **in urine** | -0.02 (0.01, 33.3%) | 0.02 (0.01, 27.1%) | -0.03 (0.02, 20.5%) | 0.03 (0.02, 57%) |
| **Urea** | -0.06 (0.02, 85.8%) | 0.02 (0.02, 8.77%) | -0.07 (0.02, 88.6%) | 0.03 (0.01, 3.4%) |
| **CKD** | -0.32 (0.11, 90.1%) | 0.13 (0.04, 1.48%) | -0.38 (0.12, 91.1%) | 0.11 (0.04, 3.08%) |
| **T2D** | -0.06 (0.01, 0.98%) | 0.17 (0.06, 85.7%) | -0.02 (0.01, 0.05%) | 0.25 (0.14, 86.1%) |
| **Hypertension** | -0.04 (0.02, 25.2%) | 0.06 (0.04, 53.6%) | -0.04 (0.02, 6.69%) | 0.06 (0.04, 76%) |
| **Age * CKD** | -0.10 (0.04, 44.8%) | - | -0.09 (0.02, 29.8%) | 0.10 (0.03, 10.8%) |
| **Age * T2D** | -0.08 (0.03, 82%) | 0.06 (0.01, 0.73%) | -0.08 (0.04, 52.4%) | 0.002 (0.001, 0.09%) |
| **Age * Hypertension** | -0.03 (0.01, 40.3%) | 0.04 (0.004, 0.3%) | -0.03 (0.01, 30.4%) | 0.003 (0.003, 0.09%) |
| **BMI * T2D** | -0.08 (0.04, 67%) | 0.06 (0.02, 7.51%) | -0.06 (0.02, 24.1%) | 0.08 (0.02, 2.33%) |
| **WHR * T2D** | -0.08 (0.03, 0.64%) | 0.07 (0.02, 15.4%) | -0.08 (0.03, 28.9%) | 0.02 (0.01, 0.05%) |

**Table S4.** Significance areas for covariates in the MUR model between the anthropometric covariates in the model for the left kidney for each gender (18,855 males and 20,013 females). The total area has been split into areas of positive and negative associations. The standardised regression coefficients ($\hat{\beta}$) are presented as median (interquartile range - IQR) across all vertices of the left kidney surface and the significance areas as a percentage (%) of the vertices with statistically significant associations.

| **Right Kidney** | **Male** | | **Female** | |
| --- | --- | --- | --- | --- |
|  | $\hat{\beta}<0$ | $\hat{\beta}>0$ | $\hat{\beta}<0$ | $\hat{\beta}>0$ |
|  | Median Beta coefficients (IQR, Significance area (%)) | | | |
| **Age** | -0.08 (0.06, 79.4%) | 0.05 (0.04, 8.36%) | -0.08 (0.05, 74.1%) | 0.04 (0.04, 15.7%) |
| **BMI** | -0.16 (0.05, 12%) | 0.28 (0.13, 87.8%) | -0.14 (0.04, 12.8%) | 0.25 (0.08, 87%) |
| **WHR** | -0.05 (0.05, 51.2%) | 0.04 (0.04, 33.7%) | -0.06 (0.05, 15.1%) | 0.04 (0.04, 71.7%) |
| **Alcohol intake frequency** | -0.05 (0.02, 12%) | 0.07 (0.02, 86.5%) | -0.06 (0.04 12.1%) | 0.10 (0.04, 84.5%) |
| **Smoking status** | -0.08 (0.02, 2.3%) | 0.13 (0.09, 92.4%) | -0.09 (0.07,1.13%) | 0.16 (0.08, 88.1%) |
| **Ibuprofen medication** | -0.02 (0.01, 26%) | 0.06 (0.03, 76.8%) | -0.03 (0.03,0.74%) | 0.04 (0.02, 72.3%) |
| **Sodium**  **in urine** | -0.02 (0.01, 21.8%) | 0.02 (0.01, 45.3%) | -0.02 (0.02, 34.1%) | 0.02 (0.01, 40.8%) |
| **Urea** | -0.06 (0.01, 87.7%) | 0.02 (0.01, 7%) | -0.06 (0.02, 94.4%) | 0.02 (0.01, 3.26%) |
| **CKD** | -0.36 (0.15, 92.9%) | 0.12 (0.07, 1.9%) | -0.30 (0.12, 95.2%) | 0.11 (0.03, 1.99%) |
| **T2D** | -0.06 (0.02, 5.32%) | 0.20 (0.08, 88%) | -0.09 (0.04, 1.51%) | 0.23 (0.08, 85.2%) |
| **Hypertension** | -0.05 (0.03, 15.4%) | 0.08 (0.05, 72.5%) | -0.03 (0.02, 6.35%) | 0.06 (0.04, 85.7%) |
| **Age * CKD** | -0.11 (0.03, 18.5%) | 0.01 (0.01, 0.22%) | -0.08 (0.04, 65.8%) | 0.09 (0.03, 8.22%) |
| **Age * T2D** | -0.07 (0.04, 85.6%) | 0.07 (0.05, 0.72%) | -0.08 (0.04, 42.4%) | 0.01 (0.001, 0.02%) |
| **Age * Hypertension** | -0.03 (0.01, 49.6%) | 0.04 (0.03, 1.27%) | - | - |
| **BMI * T2D** | -0.08 (0.05, 80.5%) | 0.08 (0.03, 12%) | -0.09 (0.05, 43.2%) | 0.08 (0.03, 17.9%) |
| **WHR * T2D** | -0.07 (0.03, 1.49%) | 0.08 (0.03, 22.6%) | -0.07 (0.02, 4.65%) | 0.10 (0.02, 7.43%) |

**Table S5.** Significance areas for covariates in the MUR model between the anthropometric covariates (N=38,868) in the model for the right kidney for each gender (18,855 males and 20,013 females). The total area has been split into areas of positive and negative associations. The standardised regression coefficients ($\hat{\beta}$) are presented as median (interquartile range - IQR) across all vertices of the left kidney surface and the significance areas as a percentage (%) of the vertices with statistically significant associations.

Supplementary Figures

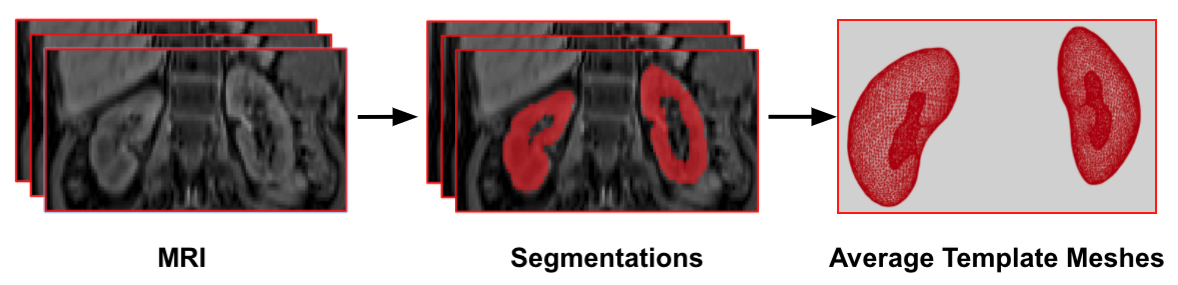

**Figure S1.** Average template mesh construction. Dixon MRI volumes from UK Biobank abdominal protocol (left) are used to produce subject-specific 3D kidneys segmentations (middle), then images are registered to a common space and combined to produce average kidneys template meshes.

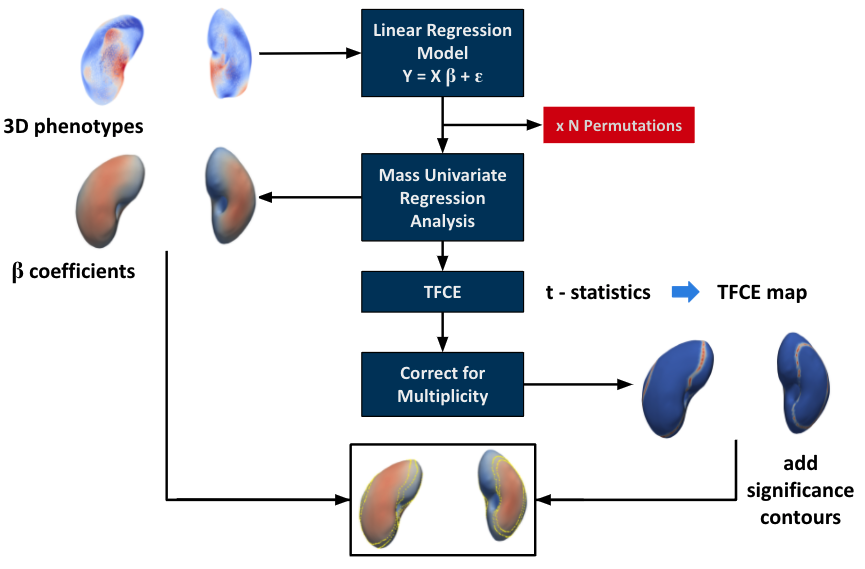

**Figure S2.** Flow diagram for the mass univariate regression (MUR) analysis of three-dimensional phenotypes. The phenotypes are used to construct the linear regression model. MUR analysis produces parameter estimates ($\hat{\beta}$) and their null distribution via permutation. Threshold free cluster enhancement (TFCE) is applied to the $t$-statistics from the regression analysis to produce a significance threshold. The associated TFCE-derived $p$- values are corrected for multiple comparisons and mapped onto the kidney’s mesh for visualisation. This diagram was modified from [[4]](https://paperpile.com/c/KvRrXW/GzMs).

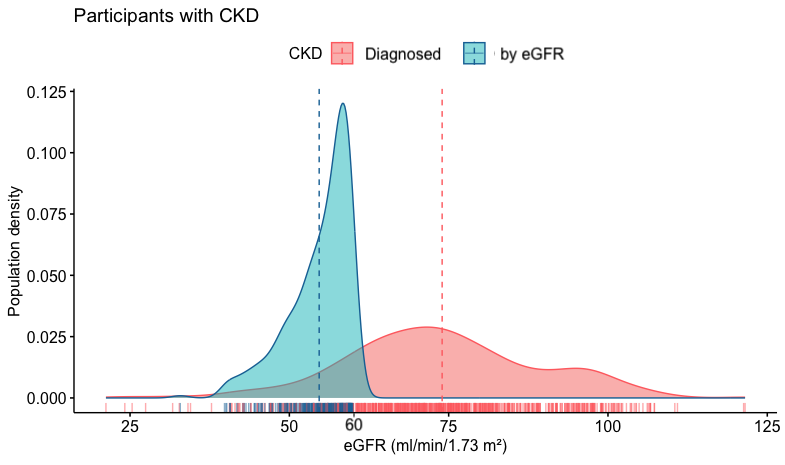

**Figure S3.** Density plots showing the participants with CKD that are diagnosed by doctor, shown in red (N =793) and selected by eGFR below 60 ml/min/1.73 m², shown in blue (N=466), across eGFR levels. Means for each CKD are shown in dashed lines.

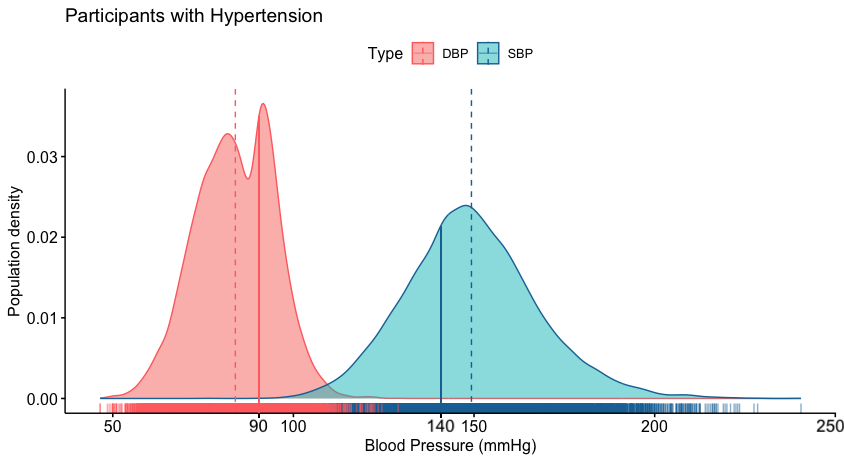

**Figure S4.** Density plots showing the participants with hypertension (N=14,113) across blood pressure readings. The thresholds applied are for systolic blood pressure (SBP) ≥ 140 mmHg shown in blue solid line and diastolic blood pressure (DBP) ≥ 90 mmHg shown in red solid line. Means for each blood pressure reading are shown in dashed lines.

**Left Kidney**

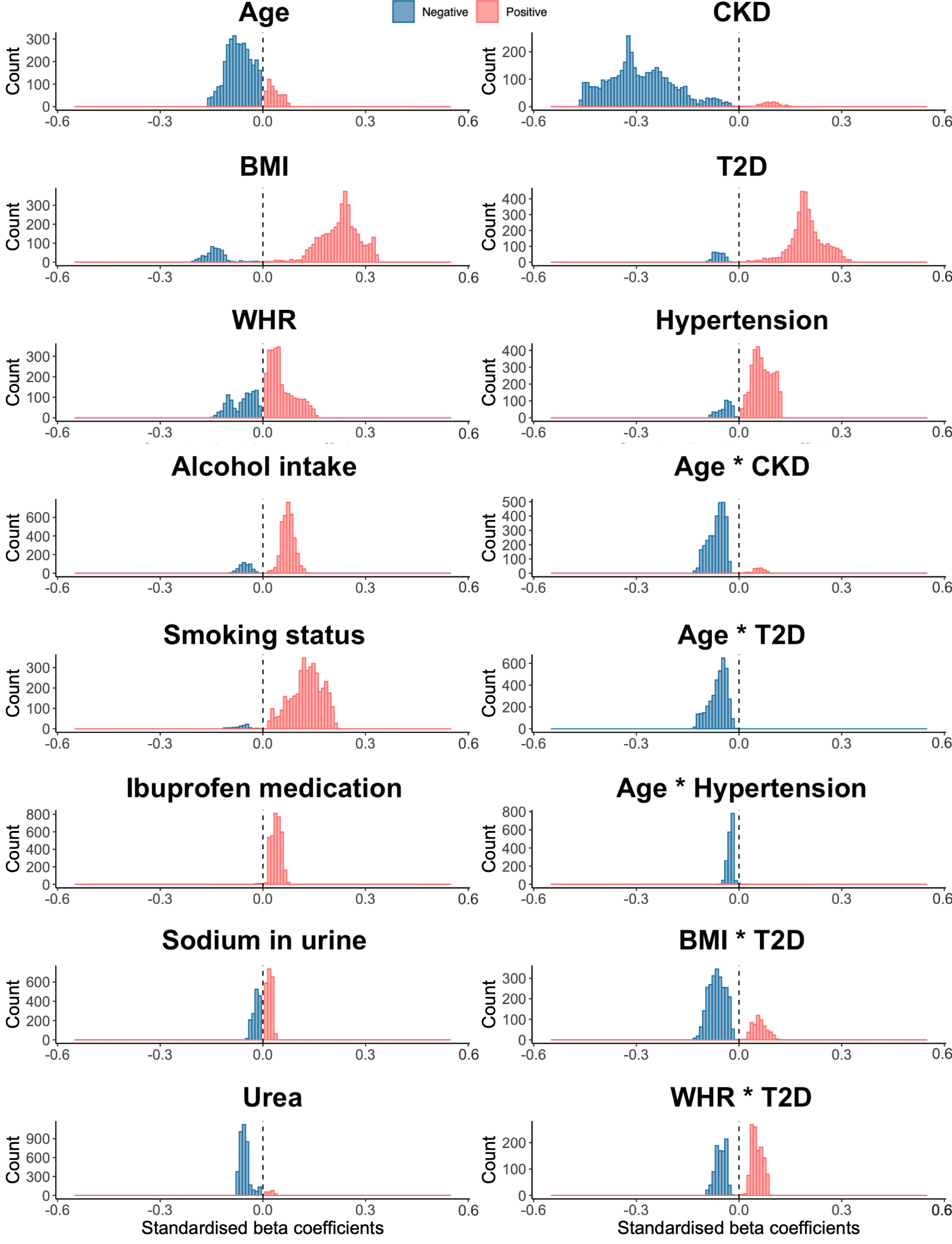

**Figure S5.** Histograms showing the statistically significant regression coefficients across the vertices (~4,000) of the left kidney for each covariate in the model on the full cohort (N=38,868) with positive associations in red and negative associations in blue. Beta coefficients are provided with units in standard deviations for each covariate.

**Right Kidney**

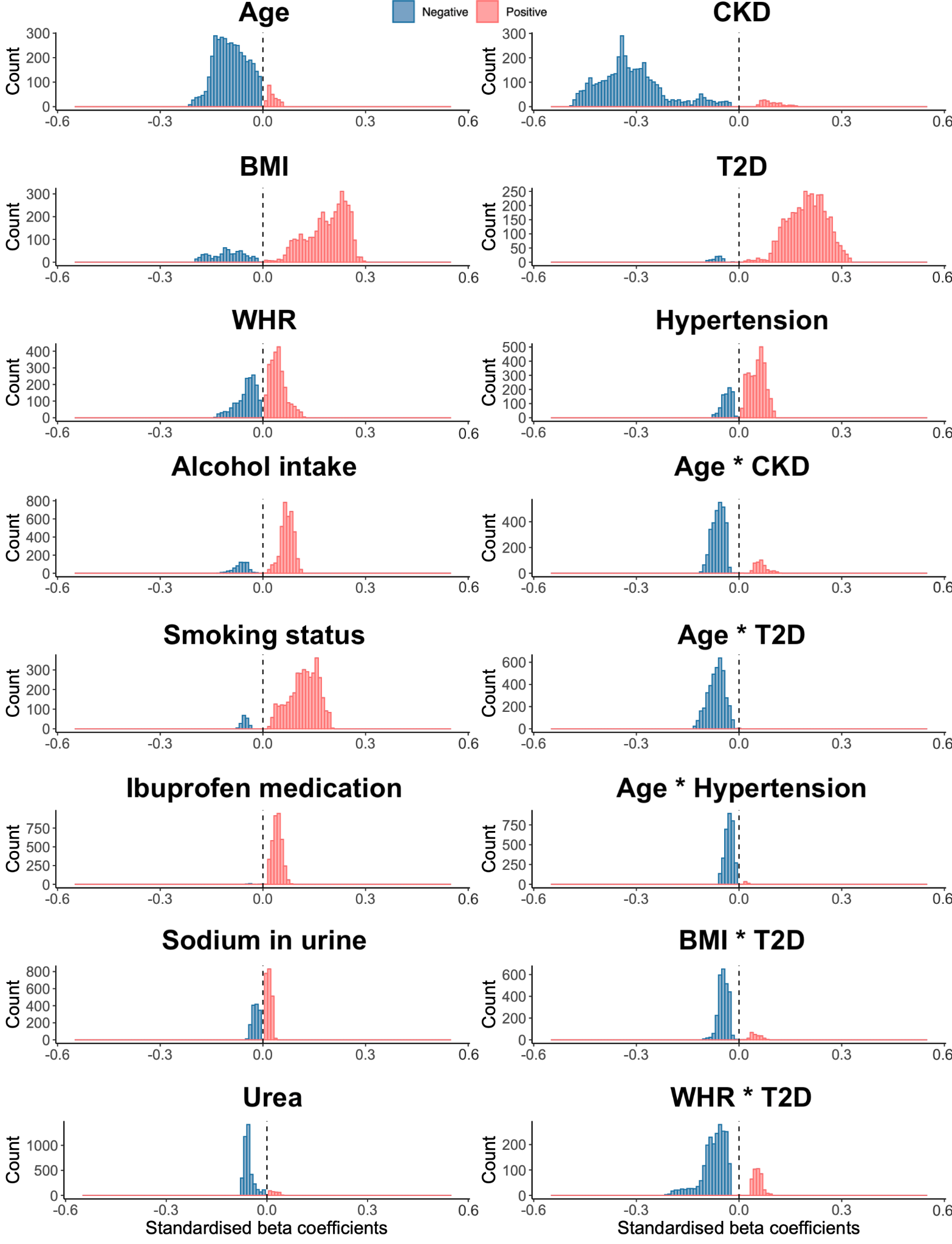

**Figure S6.** Histograms showing the statistically significant regression coefficients across the vertices (~4,000) of the right kidney for each covariate in the model on the full cohort (N=38,868) with positive associations in red, negative associations in blue. Beta coefficients are provided with units in standard deviations for each covariate.

**Left Kidney**

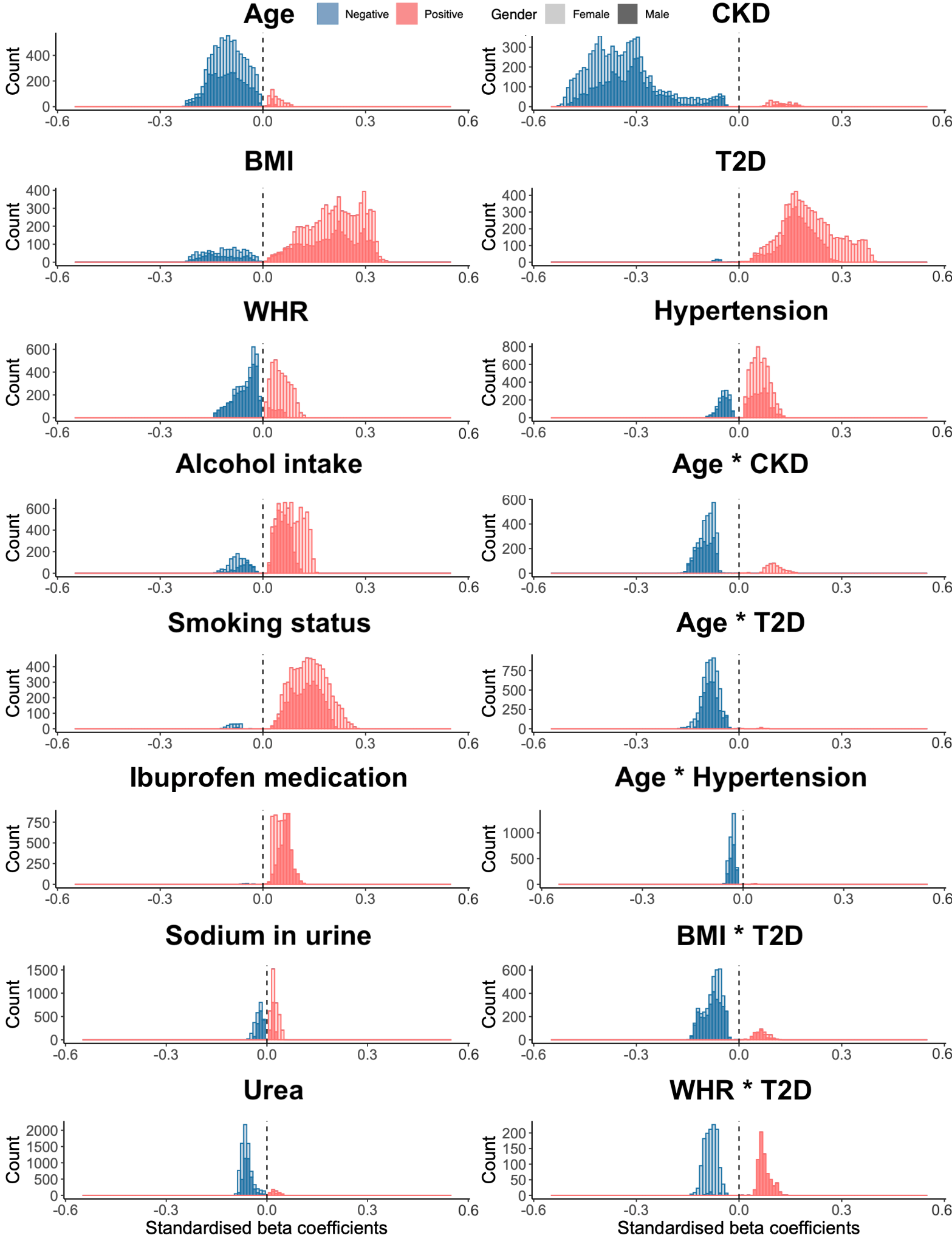

**Figure S7.** Histograms showing the statistically significant regression coefficients across the vertices (~4,000) of the left kidney for each covariate in the model on the full cohort (N=38,868) separated by gender with positive associations in red, negative associations in blue, female (N=20,013) in light colour and male (N=18,855) in darker colour. Beta coefficients are provided with units in standard deviations for each covariate.

**Right Kidney**

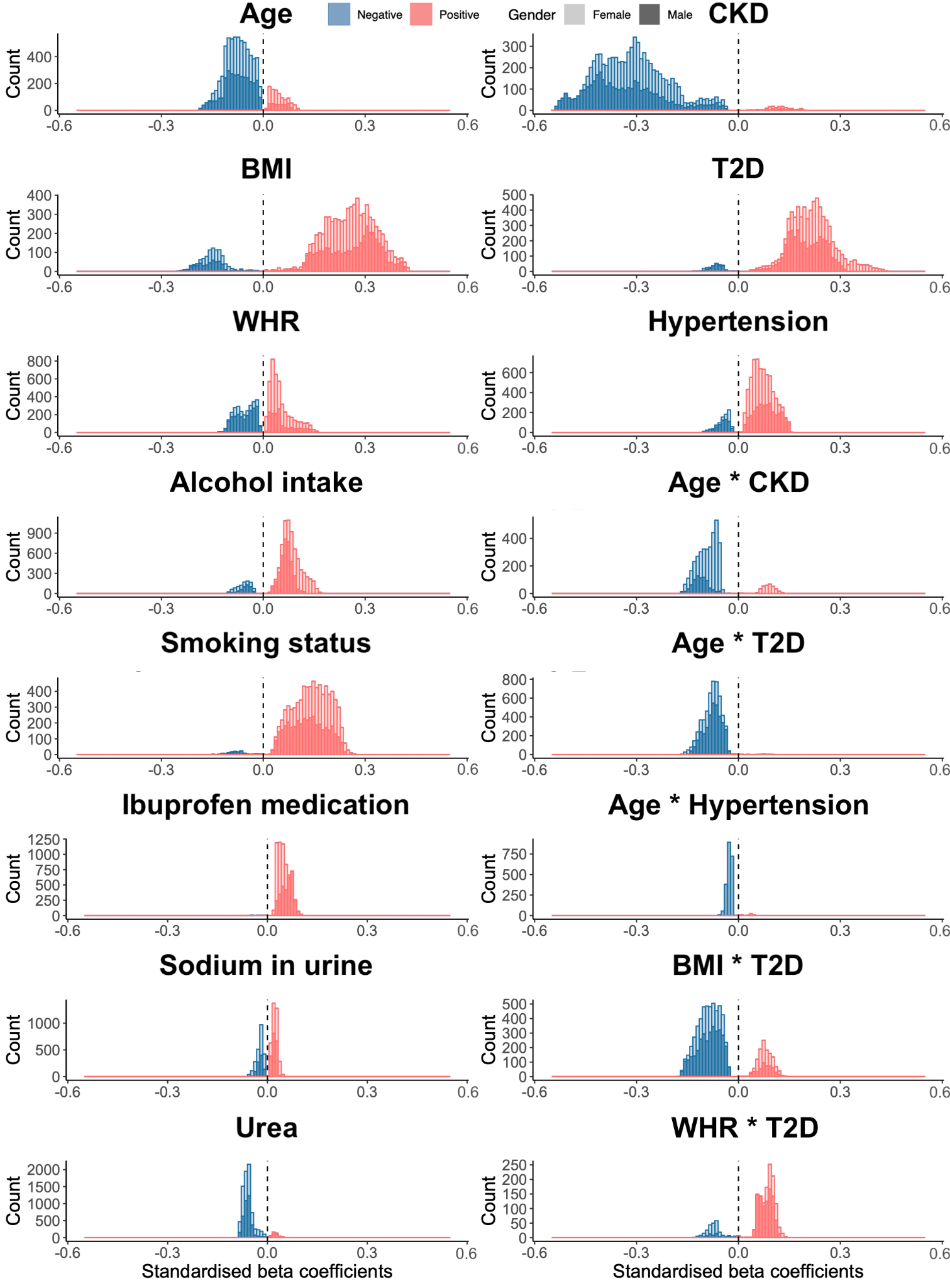

**Figure S8.** Histograms showing the statistically significant regression coefficients across the vertices (~4,000) of the right kidney for each covariate in the model on the full cohort (N=38,868) separated by gender with positive associations in red, negative associations in blue, female (N=20,013) in light colour and male (N=18,855) in darker colour. Beta coefficients are provided with units in standard deviations for each covariate.

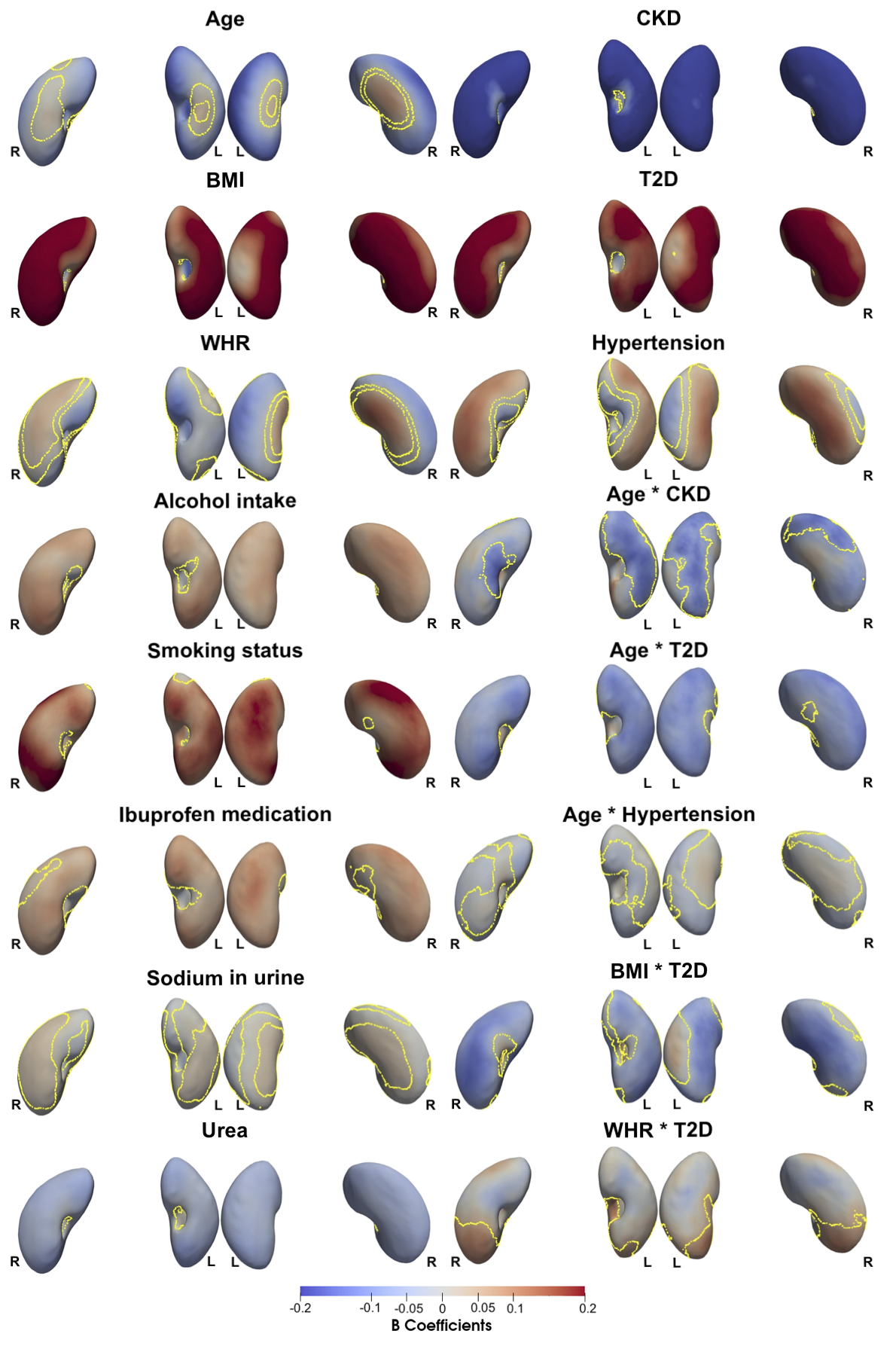

**Figure S9.** Three-dimensional statistical parametric maps (SPMs) of kidney morphology, projections are anterior (left plots) and posterior (right plots) views for both left (L) and right (R) kidneys in both anterior (left) and posterior (right) views. The SPMs show the local strength of association for each covariate in the model with S2S distances on the male cohort (N=18,855). Yellow contour lines indicate the boundary between statistically significant regions (p < 0.05) after correction for multiple testing, with positive associations in bright red and negative associations in bright blue. Regression coefficients are shown with units in standard deviations for each covariate.

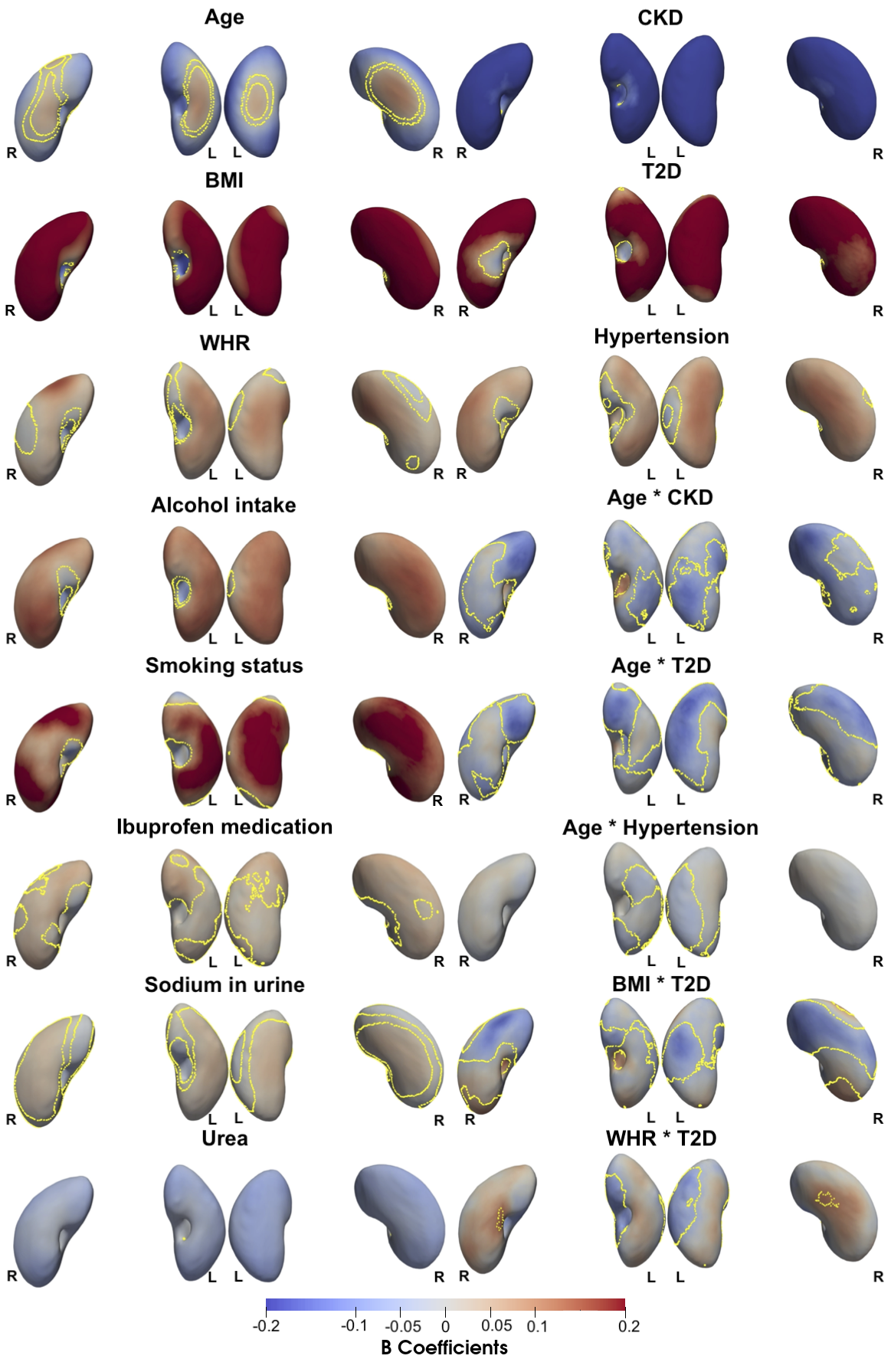

**Figure S10** Three-dimensional statistical parametric maps (SPMs) of kidney morphology, projections are anterior (left plots) and posterior (right plots) views for both left (L) and right (R) kidneys in both anterior (left) and posterior (right) views. The SPMs show the local strength of association for each covariate in the model with S2S distances on the female cohort (N=20,013). Yellow contour lines indicate the boundary between statistically significant regions (p < 0.05) after correction for multiple testing, with positive associations in bright red and negative associations in bright blue. Regression coefficients are shown with units in standard deviations for each covariate.

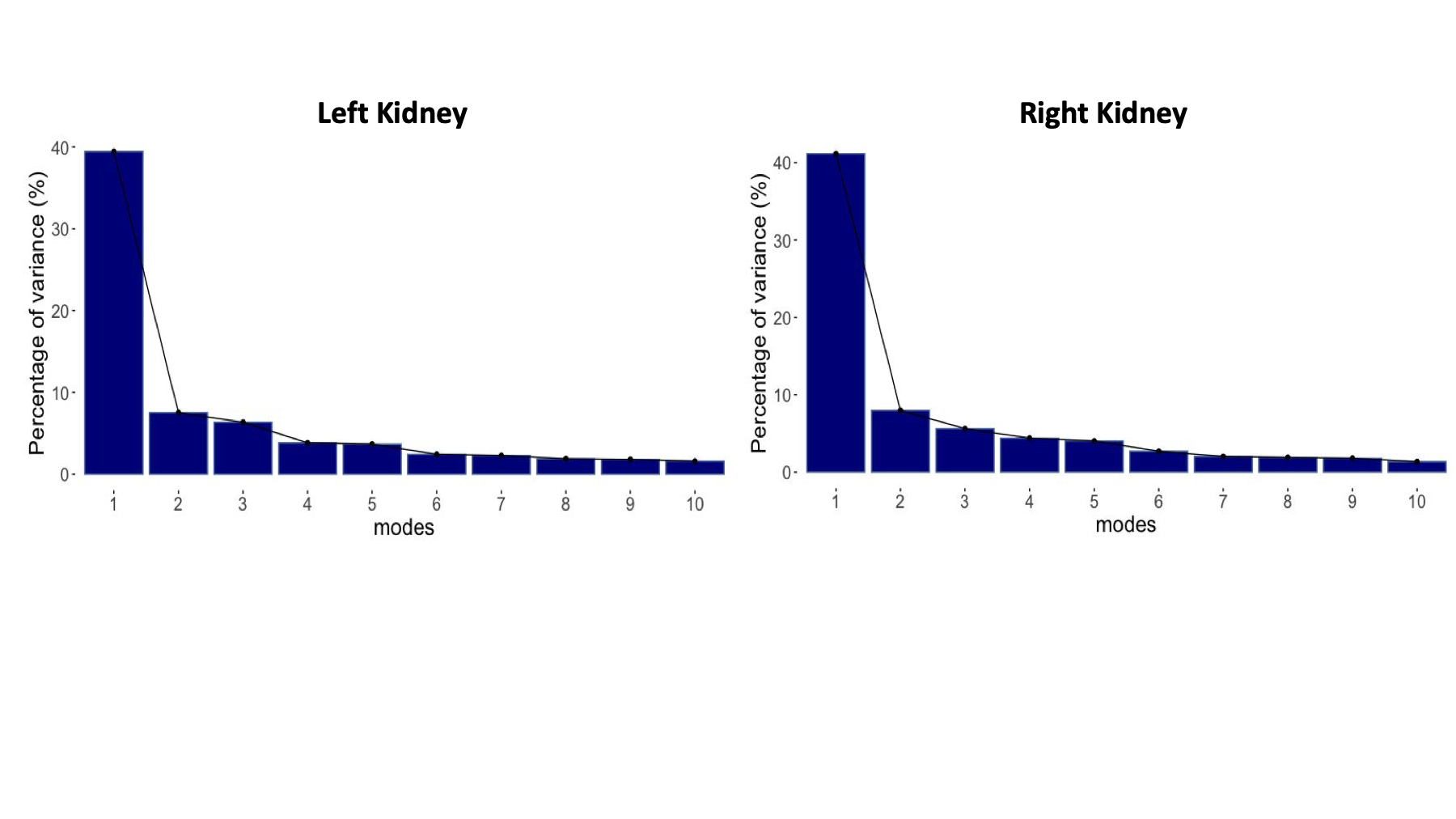

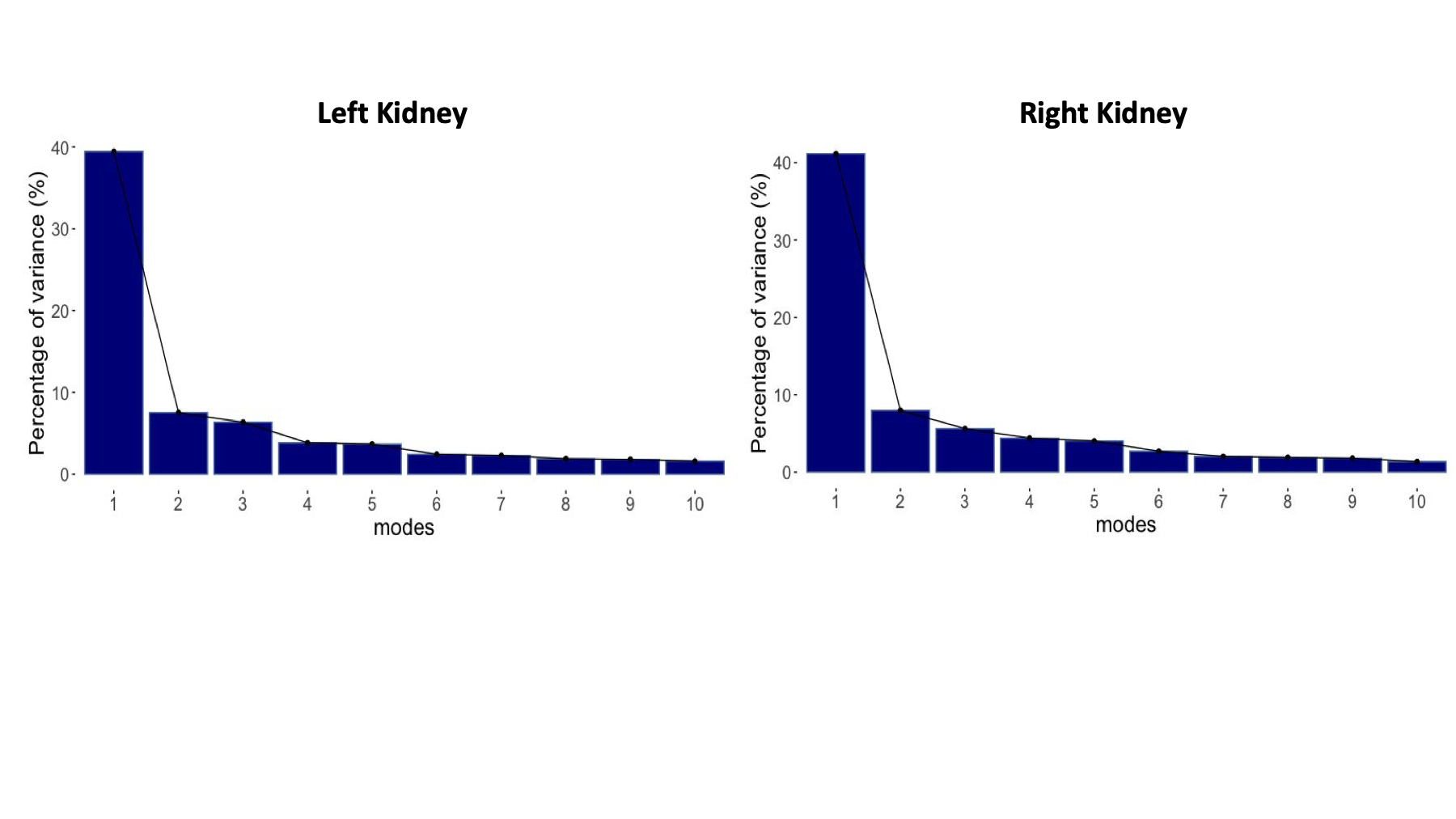
**Figure S11.** The percentage of shape variation explained by the first ten modes of PCA for the kidneys of the full cohort (N=38,868).

Supplementary Videos

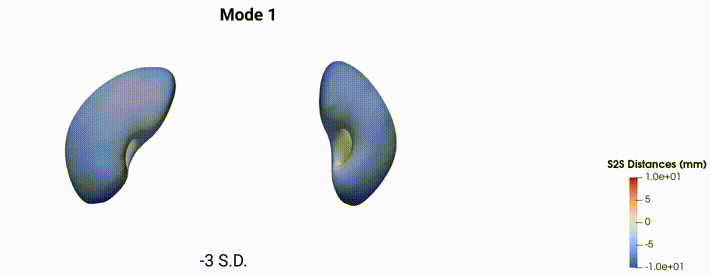

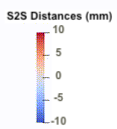

**Video S1.** The first 4 modes of shape variation for the kidneys of the full cohort (N=38,868). The mean shape and the shape at the +/- 3 standard deviations are displayed for each mode showing the S2S distance change in mm. The right kidney is shown on the left side and the left kidney on the right side of the video.
